# A multi-phenotype genome-wide association study of clades causing tuberculosis in a Ghanaian- and South African cohort

**DOI:** 10.1101/2020.07.27.20162925

**Authors:** Stephanie J. Müller, Haiko Schurz, Gerard Tromp, Gian D. van der Spuy, Eileen G. Hoal, Paul D. van Helden, Ellis Owusu-Dabo, Christian G. Meyer, Thorsten Thye, Stefan Niemann, Robin M. Warren, Elizabeth Streicher, Marlo Möller, Craig Kinnear

## Abstract

Despite decades of research and advancements in diagnostics and treatment, tuberculosis remains a major public health concern, particularly in low- and middle-income countries. New bioinformatics and computational methods are needed to interrogate the intersection of host- and bacterial genomes and identify novel targets for anti-tuberculosis drugs. Host genotype datum and paired infecting bacterial isolate information were analysed for associations using a multinomial logistic regression framework implemented in SNPTest. Two geographically distinct cohorts were evaluated: a cohort of 947 participants self-identifying as belonging to a five-way admixed South African population and a Ghanaian cohort consisting of 3 311 participants. We report potential associations between host genetic variants and multiple members of the *Mycobacterium tuberculosis* complex (MTBC). Although none of the variants analyzed in the South African cohort passed the GWAS cut-off for significance, 32 single nucleotide polymorphisms were identified in the Ghanaian cohort as being statistically significantly associated with risk for infection with strains of different members of the MTBC. Further analysis revealed that two of these SNPs were directly genotyped, and the rest were imputed using the 1000 Genomes Phase 3 reference panel. The availability of paired host-pathogen data is imperative for investigating strain-specific interactions between MTBC and its host. As demonstrated by this study, the implementation of a multinomial logistic regression using paired host-pathogen data may prove valuable for further research investigating the complex relationships driving infectious disease.

## 1. Introduction

Tuberculosis (TB), a disease primarily affecting the lungs, is caused by pathogenic members of the *Mycobacterium tuberculosis* complex (MTBC) such as *M. africanum* (*M africanum*) and *M. tuberculosis* (*M. tb*). Infection alone, however, is not sufficient to cause disease (Bellamy, 1998; Kinnear et al., 2017; Möller and Hoal, 2010). Each branch of the MTBC comprises several clades of specific strains with variable virulence and disease-causing mechanisms (Brites and Gagneux, 2015; Gagneux, 2012; Hoal et al., 2017). While *M. africanum* is the main cause of TB in West-African countries including Ghana, *M. tb* is responsible for TB cases in most other parts of the world (Gagneux and Small, 2007). In addition to socio-economic- and environmental factors (Pratiwi, 2016; Seddon et al., 2013), predisposing diseases (Cheng et al., 2016; Lim et al., 2017), and the genetic make-up of the human host (Chimusa et al., 2014; Kinnear et al., 2017; Oki et al., 2011; Png et al., 2012; Salie et al., 2014; Thye et al., 2012, 2010) have been shown to play pivotal roles in determining susceptibility to the disease.

Several associations between genomic loci and susceptibility to infectious diseases such as TB (Bellamy, 1998; Herb et al., 2007; Thye et al., 2010), malaria (Rockett et al., 2014) and HIV (Pastinen et al., 1998) have been reported using candidate-gene association studies, linkage studies, and genome-wide association studies (GWAS). Using these approaches, a number of genes encoding proteins of the host immune system have been associated with susceptibility to TB, including human leukocyte antigens (HLAs), *NRAMP1*, mannose binding lectin (*MBL*), IFN-gamma (*IFN*-ƴ), and Vitamin D Receptor (*VDR*) (Bellamy, 1998; Salie et al., 2014; Søborg et al., 2003; Yim and Selvaraj, 2010). While several studies have investigated the genetic association with TB (Chimusa et al., 2014; Hoal et al., 2017; Hong et al., 2017; Kinnear et al., 2017; Schurz et al., 2019a), few have investigated the association between gene variants and susceptibility to particular strains of the MTBC (McHenry et al., 2019; Omae et al., 2017).

Candidate gene studies compare allelic and genotyping frequencies of a specific genetic marker between a group of unrelated cases and controls. In a large cohort of 1 916 sputum-positive Ghanaian TB patients genotyped for the *ALOX5* g.760G>A variant, individuals who were heterozygous for the polymorphism were found to be at increased risk for developing TB (Herb et al., 2007). Furthermore, patients harboring the exonic variant (g.760A) had a greater association (OR= 1.70; [95% CI: 1.2–2.6]) with infection caused by *M. africanum* West Africa-2 (Herb et al., 2007). Modelling a recessive mode of inheritance, a protective association (OR= 0.60; [95% CI: 0.4-0.9]) was identified between the occurrence of TB and the *MBL2* G57E variant in another cohort of Ghanaian patients (Thye et al. 2011). TB patients belonging to the Ewe population were significantly more likely to be infected with *M. africanum* (OR= 3.02; [95% CI: 1.67–5.47]) and further stratification by lineage revealed that the association was strongly driven by infection with members of *M. africanum* West Africa-1 (Asante-Poku et al., 2015). *HLA* types are also known to be important in the immune response to pathogens.

In a South African candidate-gene association study of *HLA* alleles and the *M. tb* strain responsible for active TB, the *HLA-B27* allele was found to decrease risk for an additional disease episode due to a Beijing strain, following multiple episodes of disease caused by a Beijing strain (Salie et al., 2014). In addition, specific *HLA* types were found to be associated with disease caused by the different strains investigated (Salie et al., 2014). Finally, Caws and colleagues were investigated the susceptibility of the human host to different *M. tb* strains. Using a candidate-gene approach, a cohort of 237 adult Vietnamese TB patients were analysed. The authors concluded that for this cohort, individuals carrying the C allele of the toll-like receptor-2 (*TLR2*) T597C polymorphism were significantly more likely to develop TB caused by mycobacteria belonging to the East-Asian/Beijing strain family (OR = 1.57 [95% CI 1.15-2.15]) (Caws et al., 2008).

A limitation of the candidate-gene study design however, is that it requires an *a priori* hypothesis regarding which genes to target in the association analysis. To address this limitation, GWAS have become a popular alternative for identifying genetic associations with disease. Through genotyping of many common genetic variants, GWA studies enable a global interrogation of a host’s genome for associations to disease, without the limitation of predefined candidate genes (Hirschhorn and Daly, 2005). While genome-wide associations between the human host and TB have been extensively studied in several populations, the susceptibility of the host to different members of the MTBC has only in recent years gained some attention.

The first GWAS to investigate genetic susceptibility to strains of different MTBC lineages aimed to identify genome-wide associations with TB onset, stratifying a Thai cohort by infecting MTBC lineage, and the age at onset (Omae et al., 2017). The study initially attempted to identify age-related associations between five MTBC lineages and two age-stratified groups of TB participants, namely 219 young cases (under the age of 45), and 467 old cases (over the age of 45). To reduce the complexity of the association tests, the MTBC lineages were tested as one lineage versus a collective of all other lineages. After applying Bonferroni corrections, none of the genotyped single nucleotide polymorphisms (SNPs) reached genome-wide significance for association with either of the age-groups for any of the five MTBC lineages. However, when reducing the five lineages to two groups consisting only of ‘Beijing’ and ‘non-Beijing’ cases, and testing for age-related association to TB, the authors identified a single SNP on chromosome 1p13, rs1418425, reported to have a significant association to non-Beijing infected cases classified in the “old” age category (*P* = 1.58 x10^-7^; OR = 1.62 [95% C.I.: 1.35-1.93]). The authors were able to replicate the SNP in two independent cohorts, further demonstrating the importance of performing GWAS with a specific focus on pathogen lineage (Omae et al., 2017).

Another study investigated the coevolution of *M. tb* and it’s human host. The article is based on the hypothesis that longstanding coexistence between the human genome and *M. tb* lineage may reduce the risk of progressing to active TB or minimize the severity of disease. The authors investigated TB severity (as measured by the TBScore) in two cohorts from Uganda with paired *M. tb*-human DNA available to determine if interactions between *M. tb* lineage and human genetic variants exist. Although no association was found between lineage and disease severity, an interaction between a single nucleotide polymorphism (SNP) in SLC11A1 and the L4-Ugandan lineage were identified in both cohorts. In addition several IL12B polymorphisms were associated with disease severity (McHenry et al., 2019).

In order to improve our understanding of the genetic susceptibility to the MTBC clades, this study leveraged genome-wide genotyping data from the host and pathogen data to perform a genome-wide screen for clade-specific genetic associations in cohorts originating from two distinct populations.

## 2. Results

### 2.1. Participant recruitment, sample collection, and SNP genotyping

All participants recruited for the South African and Ghanaian cohorts provided blood and sputum samples for SNP genotyping, and MTBC strain identification, respectively. The South African cohort comprised 947 participants successfully genotyped for 397 337 variants, while the Ghanaian cohort consisted of 3 311 participants successfully genotyped for 783 338 variants (Table 1). Of the Ghanaian samples included in this study, 1 359 were TB cases and 69% of the participants were male (Table 1). Principal Component Analysis of the Ghanaian cohort showed contributing ethnicities from the Akan, Ga-Adangbe, Exe, and several other ethnic groups from northern Ghana (Thye et al. 2012).

**Table 1:** Summary of patient recruitment for the South African and Ghanaian cohorts.

| **Cohort** | **South Africa** | **Ghana** |
| --- | --- | --- |
| *Cases* | 853 | 1 359 |
| *Male* | 516 | 2 087 |
| *Female* | 431 | 1 224 |
| *Cases + Male* | 469 (55%) | 933 (69%) |
| *Cases + Female* | 384 (45%) | 426 (31%) |
| *Total number of participants* | 947 | 3 311 |
| *Total number of variants* | 397 337 | 7 838 |

### 2.2. Defining MTBC clades and superclades

#### 2.2.1. South Africa

The MTBC strains obtained in the South African cohort contained strains of eight of the 12 lineages, namely Beijing, CAS (represented as CAS1 in the infection database), Haarlem, Haarlem-like, Low-copy Clade (LCC), T, and Quebec (Fig. 2A). During the grouping strategy, Beijing and CAS were clustered to form the “BeijingCAS1” superclade (Fig. 2B). Similarly, Haarlem, Haarlem-like, and LCC cases were clustered to form the “HaarlemsLCC” superclade (Fig. 2B). A clade denoted as “Other” was also present in the South African cohort but does not appear on the phylogenetic tree (Fig. 1) and thus was kept as a distinct member during the grouping strategy. The T superclade was excluded from subsequent analysis due to low frequency in the cohort after clustering into superclades, leaving five superclades represented by this cohort (Fig. 2B). MTBC clade distributions were dominated by the LAM clade and closely followed by Beijing. Superclade distributions showed similar frequencies for LAM, BeijingCAS1, and HaarlemsLCC, while the Quebec and “Other” clades were the least abundant (Fig. 2B).

#### 2.2.2. Ghana

The strains obtained in the Ghanaian cohort contained 12 clade annotations obtained from spoligotyping. The afri-181 and afri-438 clades were represented by *M. africanum* on the phylogenetic tree and were subsequently grouped with EAI at the point of divergence, and named as the EAI_afri superclade (Fig. 1). Beijing and CAS were merged, as were Haarlem and X, into the “BeijingCAS’, and “HaarlemX” superclades, respectively. T and U clades were also clustered (Fig. 1). The “Ghana-2” clade was kept as a distinct superclade, while LAM and CAM were grouped based on the similarity in their spoligotyping patterns as illustrated in Stucki *et al*. 2016. This cohort thus contained 12 clades and six superclades after clustering (Fig. 2C, and Fig. 2D, respectively).

**Fig. 1.**
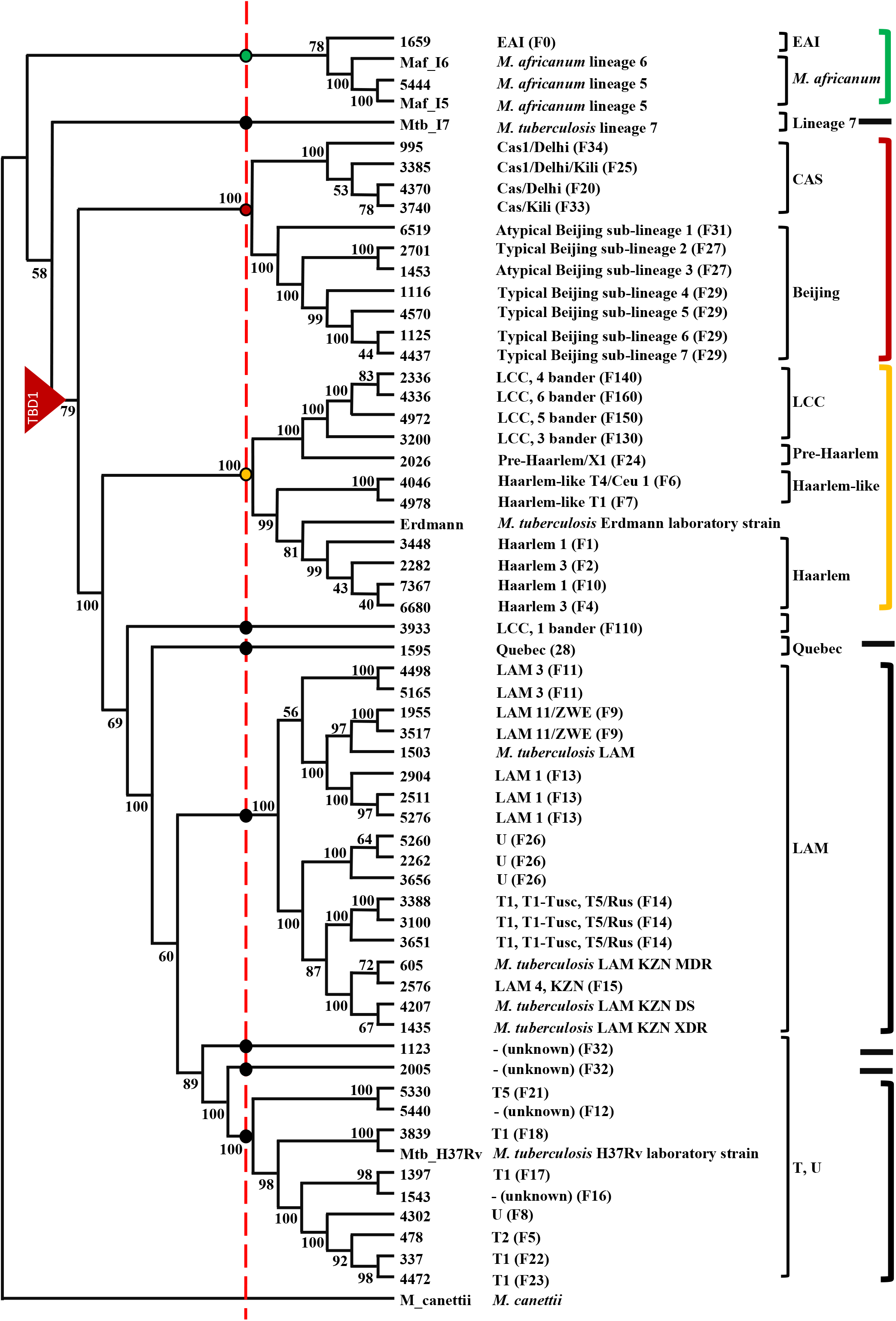
Clustering of MTBC clades on the SNP–based phylogenetic tree (adapted from original tree sourced with permission from Dippenaar 2014). MTBC clades were grouped into superclades near a point of divergence on the phylogenetic tree. Clustering reduced 12 distinct clades into seven closely related superclades. TB cases identified to be due to infection with the East African Indian (EAI) and *M. africanum* clades (green bracket), were merged into the “EAI_afri” superclade. Similarly, CAS and Beijing clades were merged into the “BeijingCAS” superclade (red bracket). The LCC, Pre-Haarlem, Haarlem-like, and Haarlem clades merged into one superclade designated as “HaarlemsLCC” (orange bracket), while the Quebec, Latin-American Mediterranean (LAM), T, and Lineage 7 clades remained unchanged due to the lack of a common progenitor on the red dotted line, were subsequently treated as individual superclades and are indicated by black brackets. The clades denoted as LCC 1 bander (F110) and ‘unknown (F32)’ were not clustered into superclades.

**Fig. 2.**
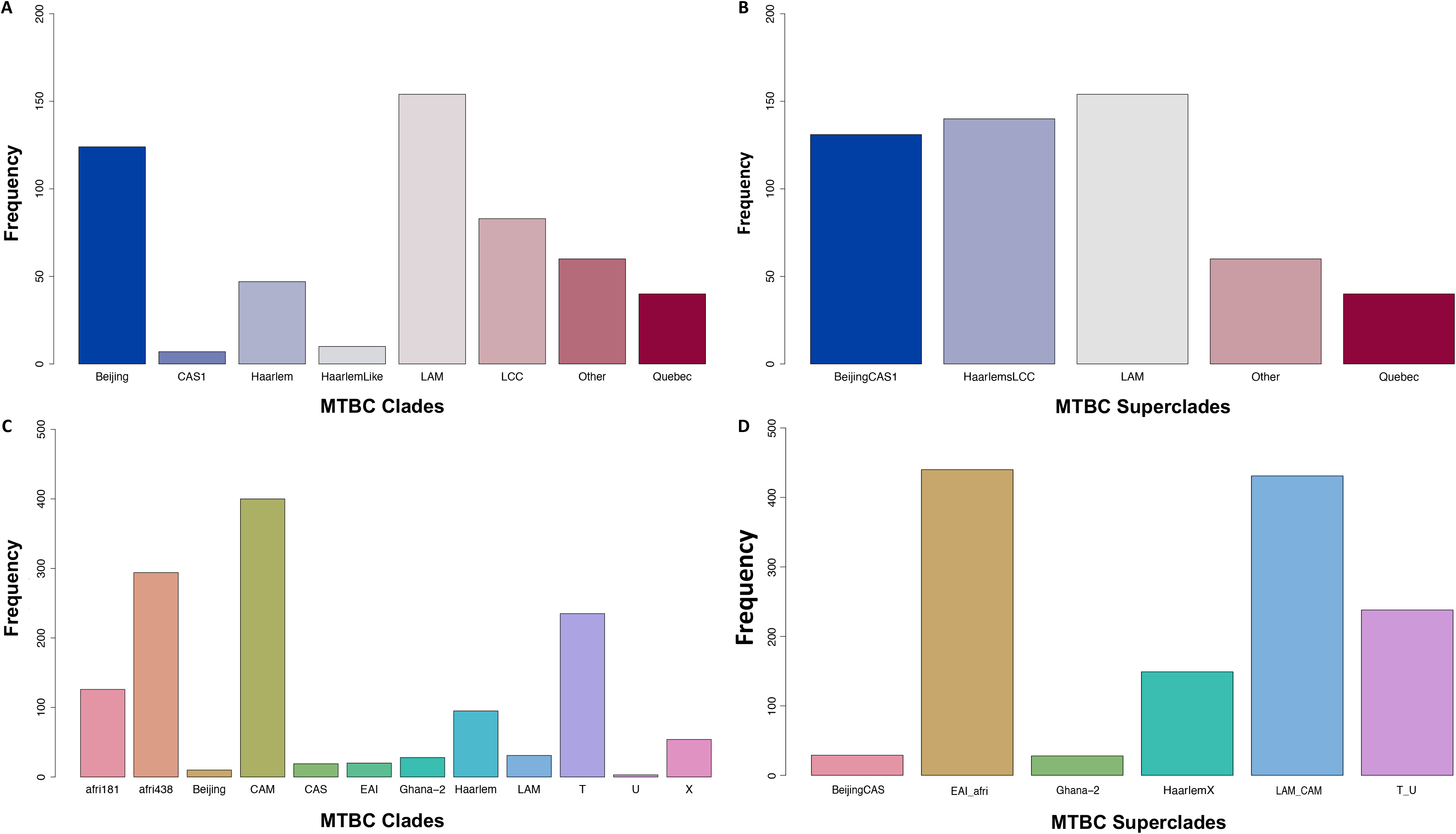
Frequency distributions of MTBC Clades and Superclades. Frequency distributions of **A:** MTBC clades and **B:** superclades for the South African cohort and **C:** clades and **D:** superclades for the Ghanaian cohort. The South African cohort was dominated by the LAM superclade, while the Ghanaian cohort was dominated by the EAI_afri and LAM_CAM superclades.

After grouping the clades into superclades, the strains of the EAI_afri and LAM_CAM superclades occurred most frequently in the cohort with both superclades having a frequency greater than 400 in the dataset (Fig. 2D). The HaarlemX, and T_U superclades had a frequency of approximately 160, and 250 in the cohort, respectively, while the BeijingCAS and Ghana-2 superclades were in least abundance (Fig. 2D).

### 2.3. Genotype data quality control, haplotype phasing, and genotype imputation

For the South African cohort, analysis using Genotype Harmonizer yielded a dataset of 947 participants with 356 165 variants. After quality control filters were applied, 919 participants and 239 612 variants remained. For the Ghanaian cohort, similar analysis yielded a dataset of 3 311 participants with 713 223 variants, and genotype QC filters resulted in a further reduction to 617 409 variants. No additional QC steps were used for the IH protocol, while the results of the additional data preparation steps for both cohorts imputed using the MIS tool are detailed in Table 2.

**Table 2:** Summary of data pre-processing on the Michigan Imputation Server.

| **Cohort** | **South Africa** |  | **Ghana** |  |
| --- | --- | --- | --- | --- |
|  | **1000G** | **CAAPA** | **1000G** | **CAAPA** |
| **Reference panel** |  |  |  |  |
| *Chromosomes* | 1–23 | 1–22 | 1–22 | 1–22 |
| **Samples** | **919** | **919** | **3 239** | **3 239** |
| *Sex undefined* | 3 | 0 | 0 | 0 |
| **SNPs** | **239 612** | **233 309** | **617 409** | **617 409** |
| *Alternative allele frequency > 0.5* | 166 | 0 | 0 | 0 |
| *Reference Overlap* | 99.49 | 97.94 | 99.55 | 97.23 |
| *Match* | 164 643 | 153 863 | 420 805 | 412 495 |
| *Allele Switch* | 74 116 | 68 912 | 176 073 | 172 432 |
| *Strand flip* | 1 | 0 | 0 | 0 |
| *Strand flip and allele switch* | 0 | 0 | 1 | 0 |
| *A/T, C/G genotypes* | 6 090 | 5 723 | 15 683 | 15 330 |
| **Filtered sites** |  |  |  |  |
| *Filter flag set* | 0 | 0 | 0 | 0 |
| *Invalid alleles* | 0 | 0 | 0 | 0 |
| *Duplicated sites* | 0 | 0 | 0 | 0 |
| *NonSNP sites* | 0 | 0 | 0 | 0 |
| *Monomorphic sites* | 0 | 0 | 0 | 0 |
| *Allele mismatch* | 54 | 8 | 2 051 | 25 |
| *SNPs call rate < 90%:* | 0 | 0 | 0 | 0 |
| *Excluded sites in total:* | 55 | 8 | 2 052 | 25 |
| **Sites remaining (before imputation)** | **239 477** | **228 498** | **612 561** | **600 257** |
| **Samples remaining** | **916** | **916** | **3 239** | **3 239** |

### 2.4. Selection of high-quality imputed genotype data

Imputation results are presented for Chromosomes 1, and X for the South African cohort, while for the Ghanaian cohort, Chromosome X data was not available, and thus the imputation of two autosomes are reported Table 3.

**Table 3:** Percentage proportion of SNPs with a quality metric greater than 0.45.

| **Cohort** | **South Africa** |  | **Ghana** |  |
| --- | --- | --- | --- | --- |
| **Chromosome** | **Chr 1** | **Chr X** | **Chr 1** | **Chr 22** |
| *IH-1000G*<sup>1</sup> | 39 | 29 | 38 | 39 |
| *MIS-1000G*<sup>2</sup> | 32 | 18 | 49 | 45 |
| *MIS-CAAPA*<sup>3</sup> | 22 | – | 56 | 50 |
| *SIS-1000G*<sup>4</sup> | 36 | 32 | 41 | 40 |
| *SIS-AGR*<sup>5</sup> | 43 | 40 | 38 | 36 |

<sup>1</sup>IH-1000G: In-House workflow using 1000G reference panel
<sup>2</sup>MIS-1000G: Michigan Imputation Server workflow using 1000G reference panel
<sup>3</sup>MIS-CAAPA: Michigan Imputation Server workflow using CAAPA reference panel
<sup>4</sup>SIS-1000G: Sanger Imputation Server workflow using 1000G reference panel
<sup>5</sup>SIS-AGR: Sanger Imputation Server workflow using AGR reference panel

SNP densities were calculated for the proportion of SNPs with a quality score metric greater than 0.45. For the South African cohort, the SIS workflow using the AGR resource imputed the highest proportion of SNPs, whereas for the Ghanaian cohort, the MIS workflow using the CAAPA resource imputed the greatest proportion of SNPs with a quality metric greater than 0.45 (Table 3). For both chromosomes 1 and X, imputation using either the 1000G or the CAAPA resource with the MIS performed the worst for the South African cohort (Schurz et al., 2019b) with the maximum median quality score only reaching 0.82 at a MAF of 50%. In comparison, the SIS-AGR workflow outperformed all other workflows, and the result correlated with the AGR imputing the highest SNP density for chromosome 1 and chromosome X (Schurz et al., 2019b).

For the South African cohort, genotype datum imputed with the AGR reference panel was selected as the dataset with the highest imputation quality across all reference panels assessed. After removal of monomorphic sites and filtering on an INFO score of 0.45, 28 566 283 SNPs for 919 samples remained. After removing the 136 related individuals identified prior to imputation, and filtering for SNP- and sample missingness, and MAF, a dataset of 7 145 406 variants for 783 participants remained. Of the 525 clade-matched samples, 445 were extracted from the dataset of samples passing QC and used in the association analysis.

For the Ghanaian cohort, despite the CAAPA resource imputing the greatest SNP density (Fig. 3D and Fig. 4D), the IH workflow using the 1000G reference panel imputed the highest quality of SNPs per MAF bin but was very closely followed by the other workflows and reference panels from the 20-30% MAF bin upwards (Fig. 5). Thus, the IH dataset, imputed with the 1000G reference panel was selected as the best dataset. After filtering monomorphic SNPs, INDELS, and variants not reaching the INFO score threshold, 25 968 622 SNPs remained for 3 239 samples. After filtering for MAF, SNP- and sample missingness, and removal of 93 related individuals identified pre-imputation, the dataset comprised of 5 275 890 variants for 1 273 clade-matched samples.

**Fig. 3.**
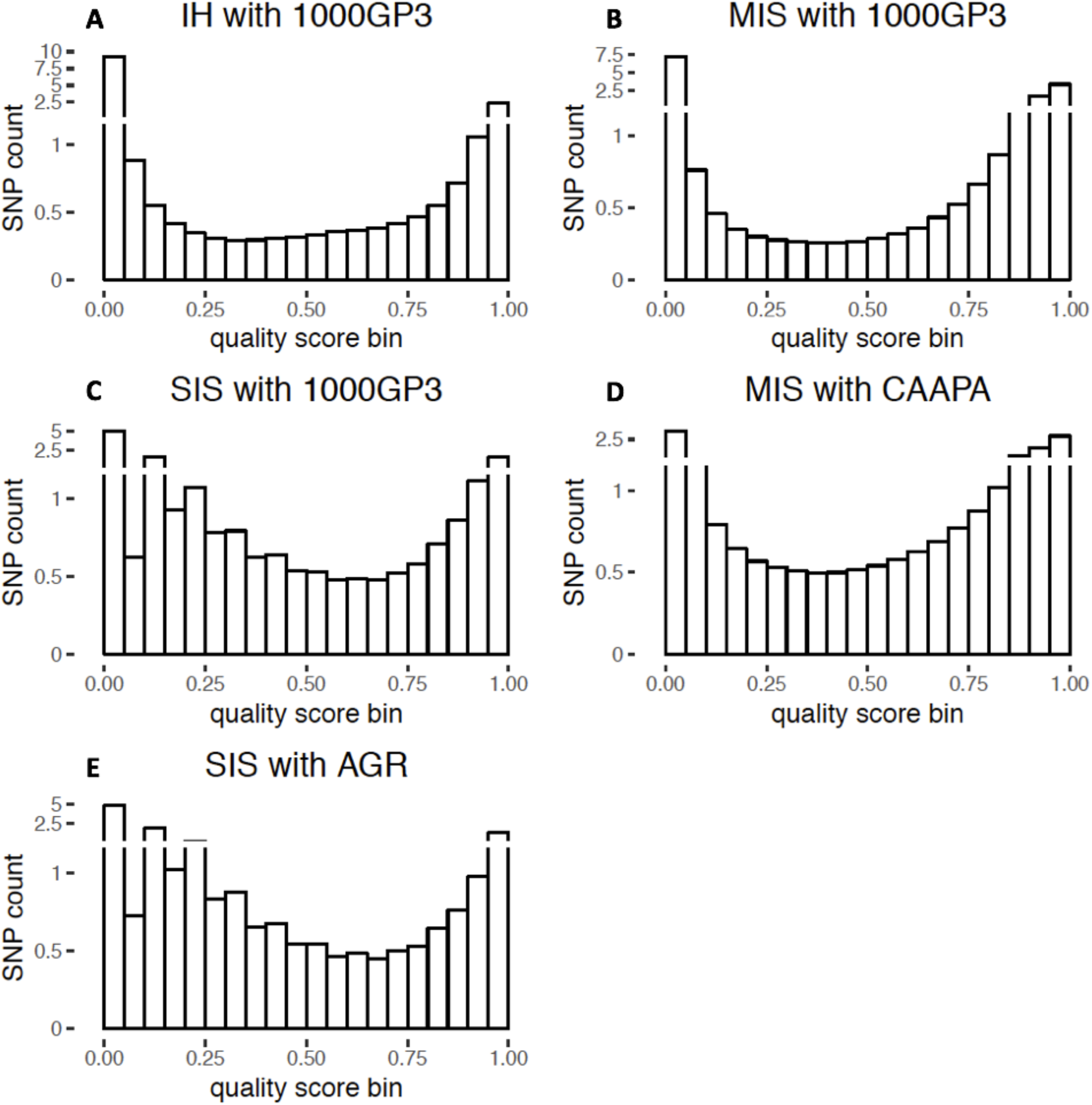
SNP density plots for Chromosome 1 of the Ghanaian cohort post-imputation using the five workflows. **A:** IH with 1000G, **B:** MIS with 1000G, **C:** SIS with 1000G, **D:** MIS with CAAPA, **E:** SIS with AGR. The MIS workflow using the CAAPA resource imputed the greatest proportion of SNPs with a quality metric greater than 0.45.

**Fig. 4.**
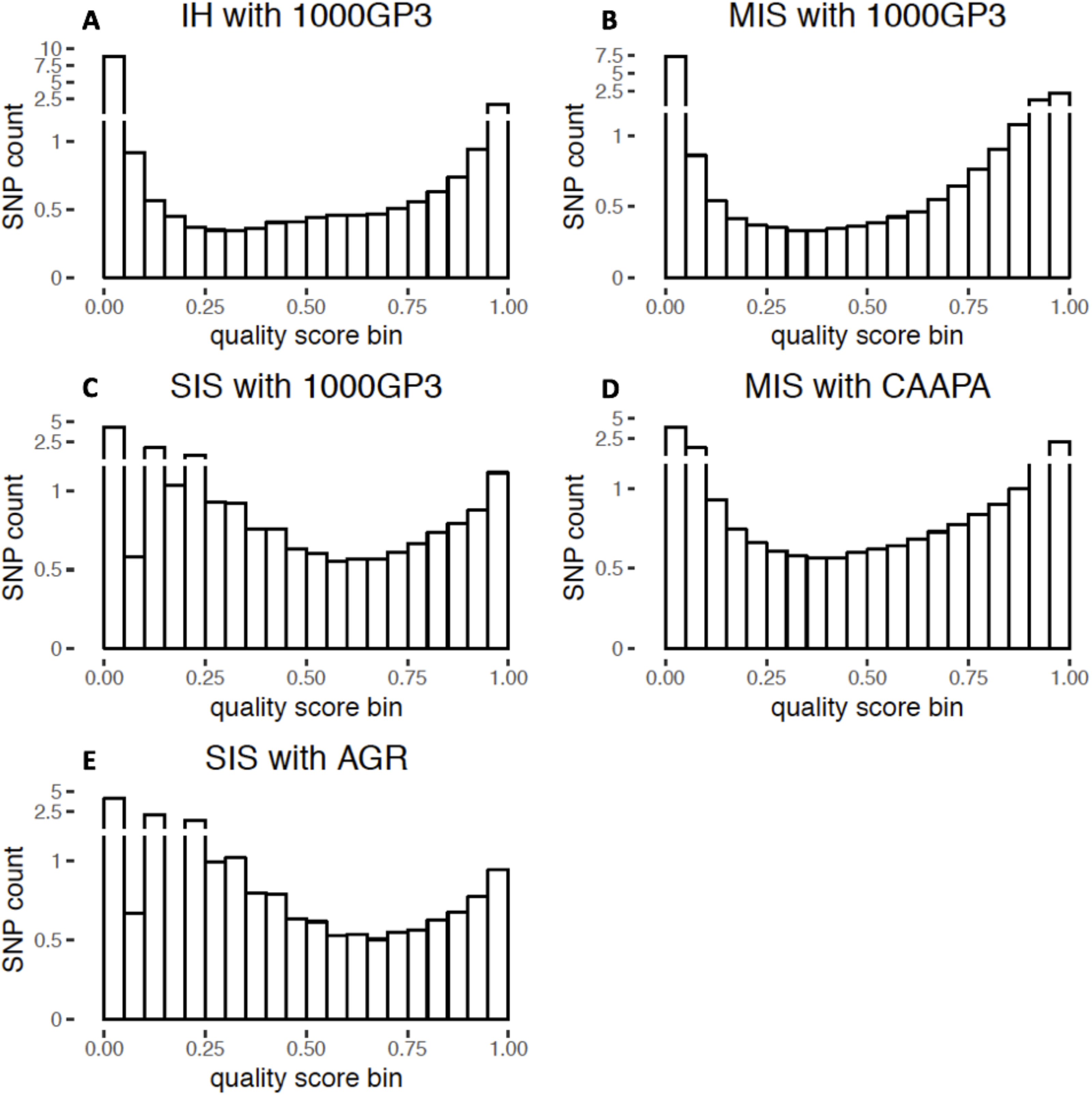
SNP density plots for Chromosome 22 of the Ghanaian cohort post-imputation using the five workflows. **A:** IH with 1000G, **B:** MIS with 1000G, **C:** SIS with 1000G, **D:** MIS with CAAPA, **E:** SIS with AGR.

**Fig. 5.**
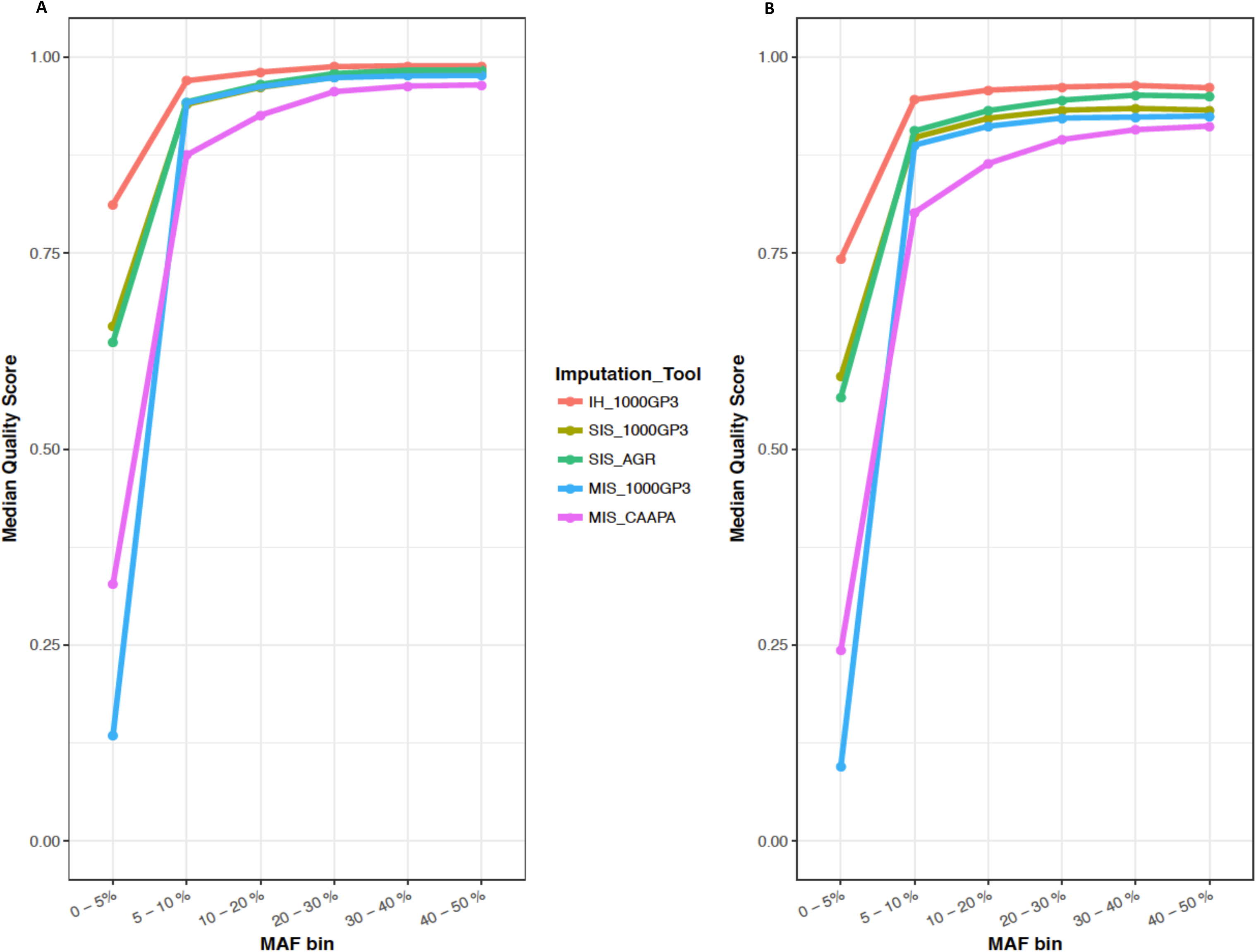
Median quality scores across MAF bins for the Ghanaian cohort, using the five protocols. **A:** Chromosome 1 **B:** Chromosome 22. For both these representative chromosomes, the IH imputation protocol using the 1000G reference panel outperformed the other four workflows.

### 2.5. Covariable data

For the South African cohort, the covariables age, sex, and ancestry proportions were available for all samples. Ancestry proportions were for the European, African, San, South-Asian, and East-Asian ancestries. Of the 445 clade-matched samples that passed the post-imputation QC, ancestry proportions were available for 357 samples as generated previously using the Affymetrix genotype datum for this cohort and ADMIXTURE software (Daya et al., 2013). For the remaining 88 samples, ancestry proportions were calculated from genotype datum generated by the MEGA array using ADMIXTURE. The East-Asian ancestry, being the smallest contributing ancestry proportion, was not included as a covariable in the analysis. Variances were calculated for each of the four remaining ancestry proportions and determined to be 0.027 (San), 0.035 (African), 0.014 (European), and 0.009 (South-Asian). As the variances were greater than the minimum cut-off of 0.001, they were included as covariables in the analysis.

For the Ghanaian cohort, age, sex, and ethnicity were included as covariables. Further examination of admixture for the Ghanaian dataset revealed that the cohort was not highly admixed, and thus ethnicity in the form of principal components was used. One sample passing QC filters did not have one of the covariables and was excluded from the dataset leaving 1 272 samples for the association analysis. The variance in the PCs provided for the cohort was calculated to be 0.0002 (PC1), 0.0003 (PC2), and 0.0004 (PC3) and therefore determined to be insufficient for inclusion in the association analysis as covariables.

### 2.6. Genome-wide Association Analyses

#### 2.6.1. South Africa

An MLR was conducted for the 445 superclade-matched South African samples using SNPTEST under an additive model. All results were reported using the LAM superclade as the baseline. Although none of the SNPs passed the GWAS cut-off for significance, 4 631 SNPs had an LRT p-value less than 0.0005 and eleven SNPs had an LRT p-value less than 1 x 10^-6^ (Table 4). Odds ratios (OR) are reported (Table 4) and standard errors of the odds ratios are shown in Fig. 6A. A single SNP, rs9389610, located on chromosome 6, had a p-value of 1.60 x 10^-7^. Individuals with the A allele of this SNP were twice as likely to be have TB due to infection with a member of the BeijingCAS1 superclade (OR: 2.19) than due to either the HaarlemsLCC (OR: 1.07) or LAM superclades. Individuals with the A allele were only slightly more at risk of being infected with a member of the ‘Other’ superclade (OR: 2.78), when compared to the BeijingCAS1 superclade (OR: 2.19), and were very unlikely to be infected with a member of the Quebec superclade (OR: 0.25).

**Table 4:** Top 11 SNPs identified by MLR to be associated with strains of different MTBC superclades in the South African cohort.

| Chr | SNP ID | Allele |  | Odd Ratio (95% C.I.) |  |  |  | LRT p-val<sup>b</sup> |
| --- | --- | --- | --- | --- | --- | --- | --- | --- |
|  |  | Ref<sup>a</sup> | Risk | BeijingCAS1 | HaarlemsLCC | Quebec | Other |  |
| 5 | rs17458866 | C | T | 0.34 (0.19–0.61) | 0.44 (0.26–0.76) | 1.99 (1.07–3.68) | 0.46 (0.23–0.95) | 10e–07 |
| 5 | rs13355101 | G | A | 0.31 (0.17–0.57) | 0.46 (0.27–0.79) | 2.00 (1.08–3.70) | 0.47 (0.23–0.96) | 6.43e–07 |
| 5 | rs12518239 | C | A | 0.29 (0.15–0.56) | 0.39 (0.22–0.70) | 1.91 (1.01–3.62) | 0.51 (0.25–1.05) | 9.41e–07 |
| 5 | rs28769614 | C | T | 0.27 (0.13–0.53) | 0.37 (0.20–0.68) | 1.92 (1.00–3.66) | 0.48 (0.23–1.01) | 3.03e–07 |
| 6 | rs9389610 | G | A | 2.19 (1.35–3.55) | 1.07 (0.64–1.76) | 0.25 (0.08–0.73) | 2.78 (1.52–5.08) | 1.60e–07 |
| 17 | rs78022196 | G | A | 1.04 (0.60–1.80) | 0.59 (0.32–1.09) | 5.31 (2.44–11.57) | 1.96 (1.03–3.73) | 5.13e–07 |
| 17 | rs72843143 | C | T | 0.98 (0.57–1.69) | 0.57 (0.31–1.04) | 4.81 (2.26–10.25) | 1.89 (1.00–3.58) | 8.18e–07 |
| 17 | rs8071332 | A | G | 0.94 (0.55–1.60) | 0.59 (0.33–1.06) | 4.77 (2.22–10.28) | 2.03 (1.08–3.81) | 6.54e–07 |
| 17 | rs10438776 | T | C | 0.99 (0.58–1.71) | 0.55 (0.30–1.01) | 4.99 (2.30–10.85) | 1.94 (1.02–3.69) | 3.93e–07 |
| 17 | rs17682747 | G | A | 0.98 (0.57–1.68) | 0.56 (0.31–1.03) | 4.85 (2.27–10.33) | 1.90 (1.01–3.58) | 5.93e–07 |
| 17 | rs7208461 | T | C | 0.94 (0.55–1.61) | 0.54 (0.30–0.98) | 4.68 (2.17–10.10) | 1.77 (0.94–3.34) | 8.64e–07 |

<sup>a</sup> Reference allele
<sup>b</sup> LAM was used as the reference superclade

**Fig. 6.**
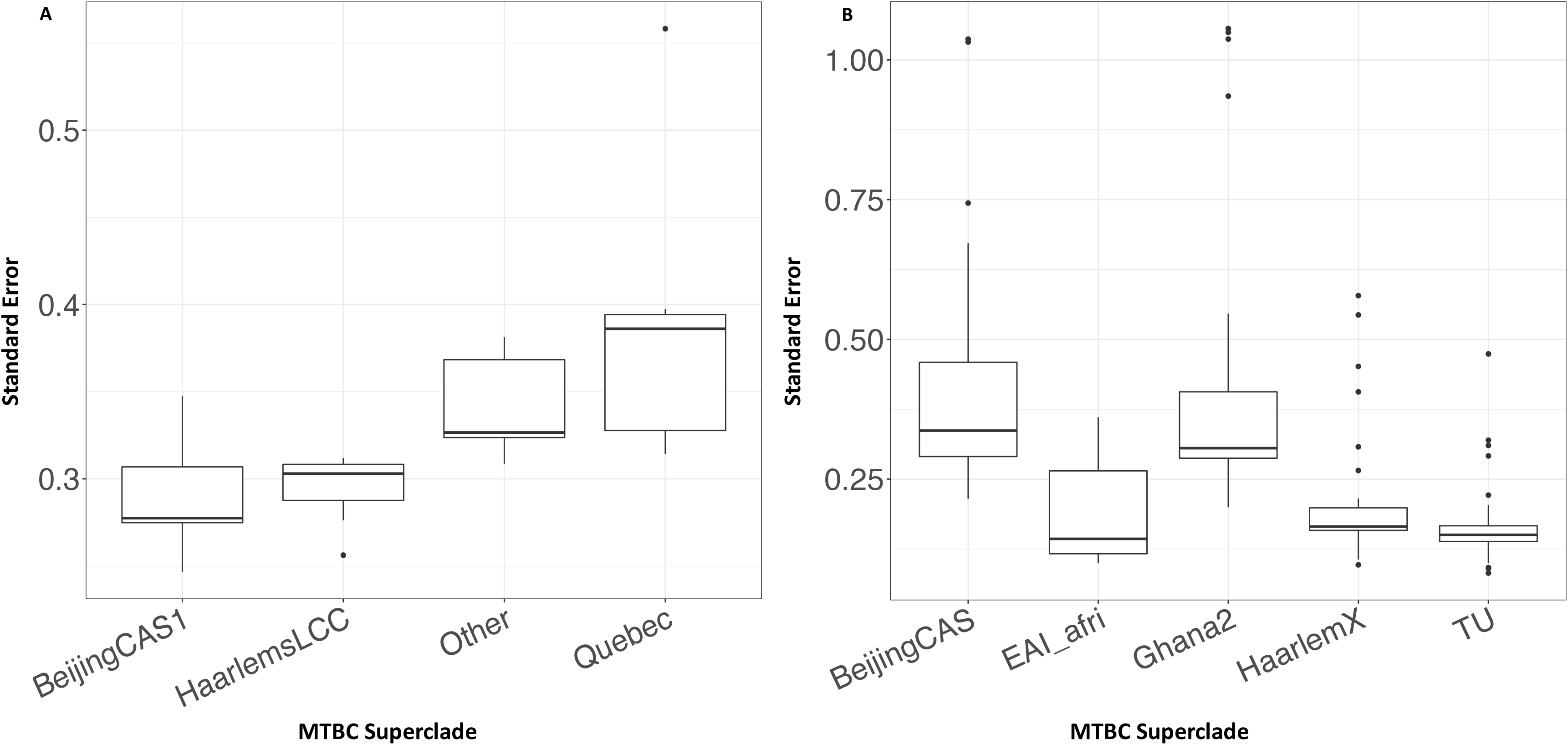
Standard errors of odds ratios calculated for each superclade against the reference LAM superclade. **A:** South African cohort **B:** Ghanaian cohort. For both cohorts, the smallest superclades had the largest variation in their standard errors.

For the four SNPs located on chromosome 5, individuals with the risk allele doubled the chances of being infected with the Quebec superclade (OR: ~2) while halving the risk of being infected with a member of the “Other” superclade (OR: ~0.5) (Table 4). Lastly, for the six SNPs located on chromosome 17, the risk allele was shown to double the risk of being infected with a member of the LAM superclade than with the HaarlemsLCC (OR: ~0.50) superclade. Individuals with the risk allele of these six SNPs were also equally at risk of being infected with a member of the BeijingCAS1 or LAM superclade and were twice as likely to be infected with the member of the “Other” superclade (OR: ~ 2) when compared to the BeijingCAS1 superclade (OR: ~ 1) (Table 4).

The VEP tool was used to retrieve gene annotations for SNPs of interest. While these SNPs are unlikely to have a direct effect on the gene expression itself, the SNP may be in linkage disequilibrium with other nearby SNPs which do have a direct effect on the gene. For the South African cohort, the most significantly associated SNP was rs9389610 (g.139039029G>A), located on chromosome 6. This SNP is an imputed SNP and its two closest directly genotyped SNPs were rs4896385 (g.139011266G>T), and rs7742202 (g.139074280A>G). The rs4896385 SNP is located in *NHSL-1*, while the rs7742202 SNP is located in *GVQW2*. Neither of these genes have been previously shown to be involved in the pathogenesis of TB. The four SNPs located on chromosome 5 (Table 4) were annotated to the StAR Related Lipid Transfer Domain Containing 4 gene (*STARD4*) using the VEP tool, while the six SNPs located on chromosome 17 were annotated to the *TANC2* gene.

#### 2.6.2. Ghana

Using SNPTEST under an additive model, an MLR was also conducted for 1 272 superclade-matched Ghanaian samples. For this cohort, all association results were reported using the LAM_CAM superclade as the baseline. In summary, a total of 32 SNPs had an LRT p-value less than 1×10^-6^ (Table 5) and were significantly associated with the MTBC superclades.

**Table 5:**
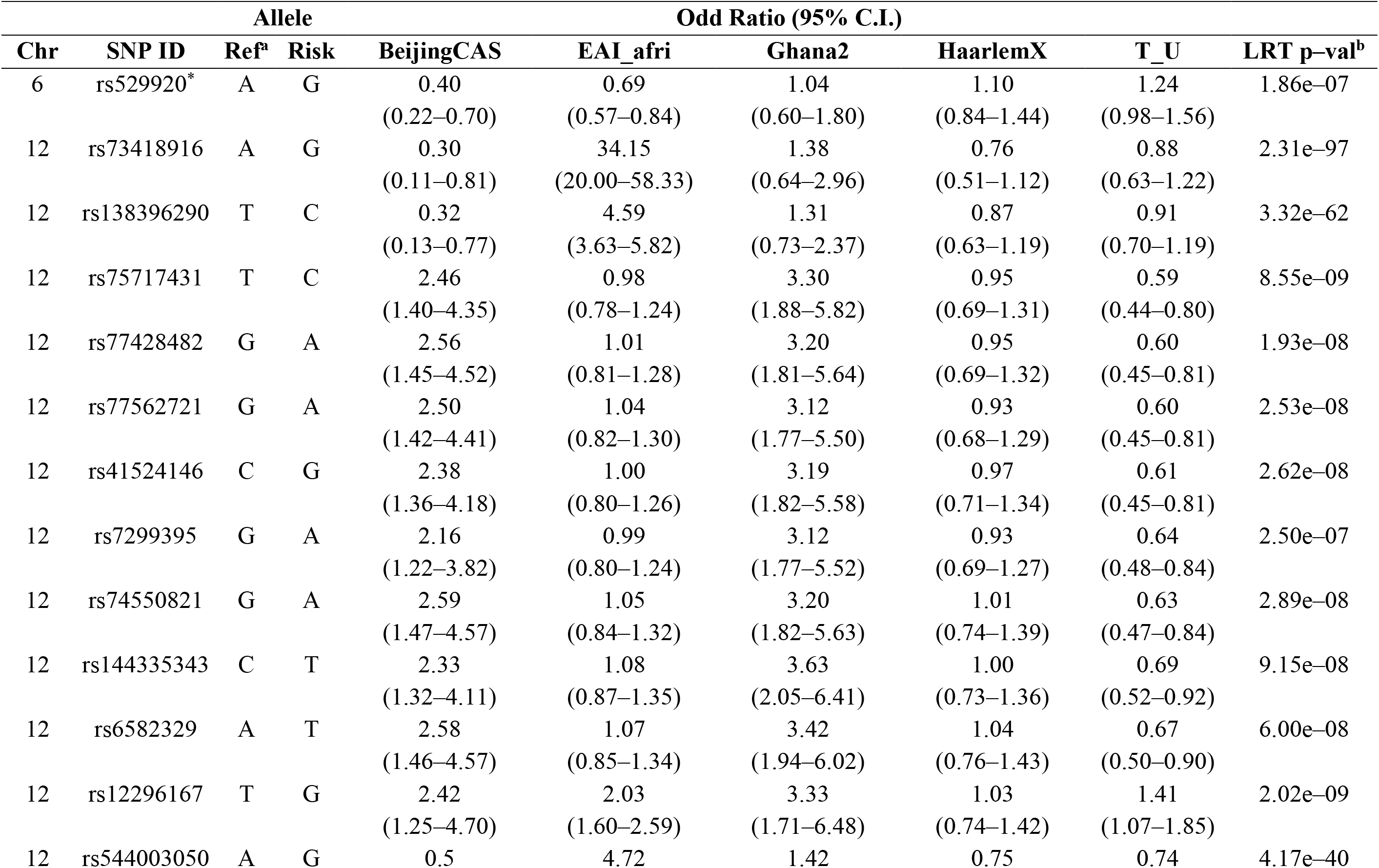
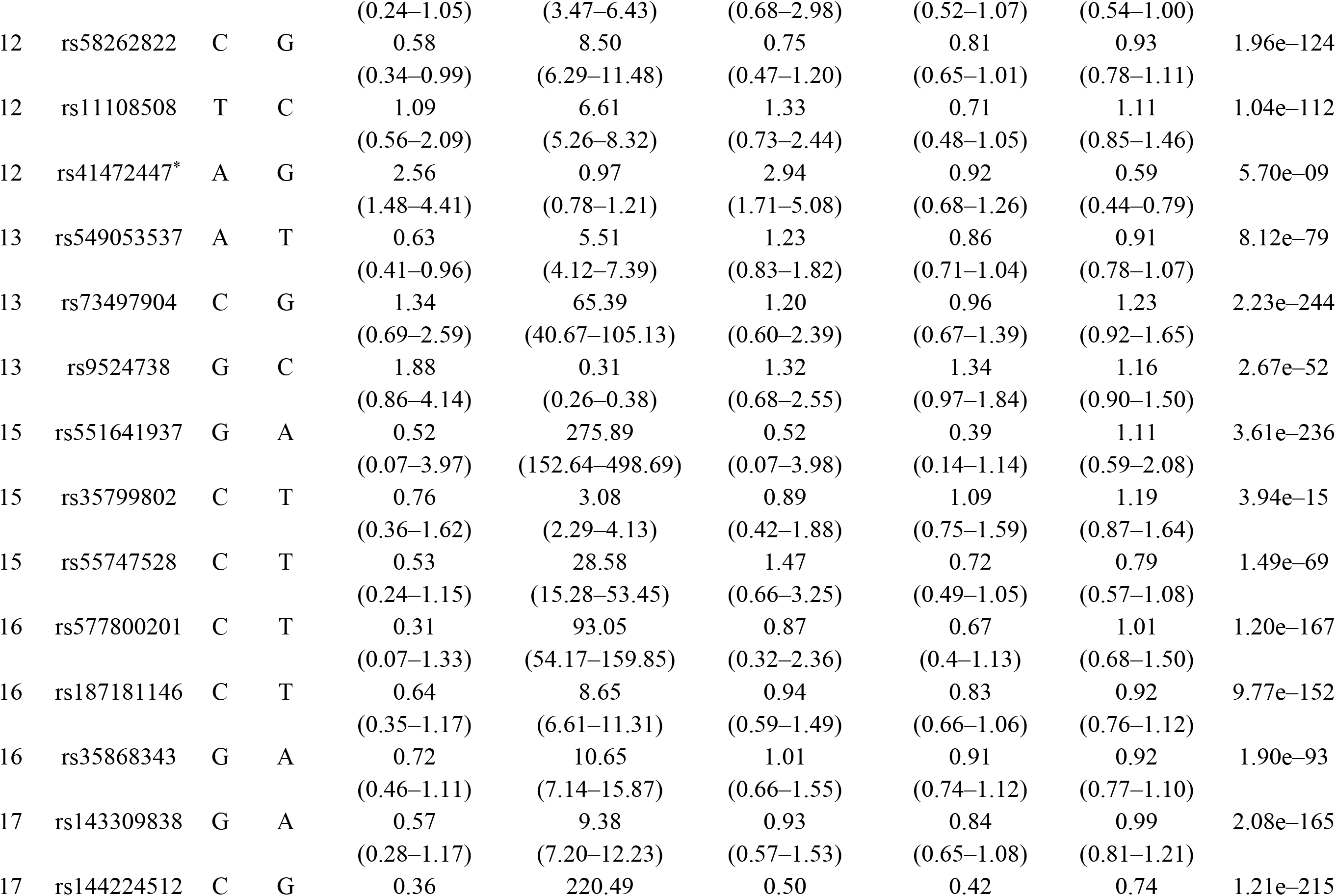
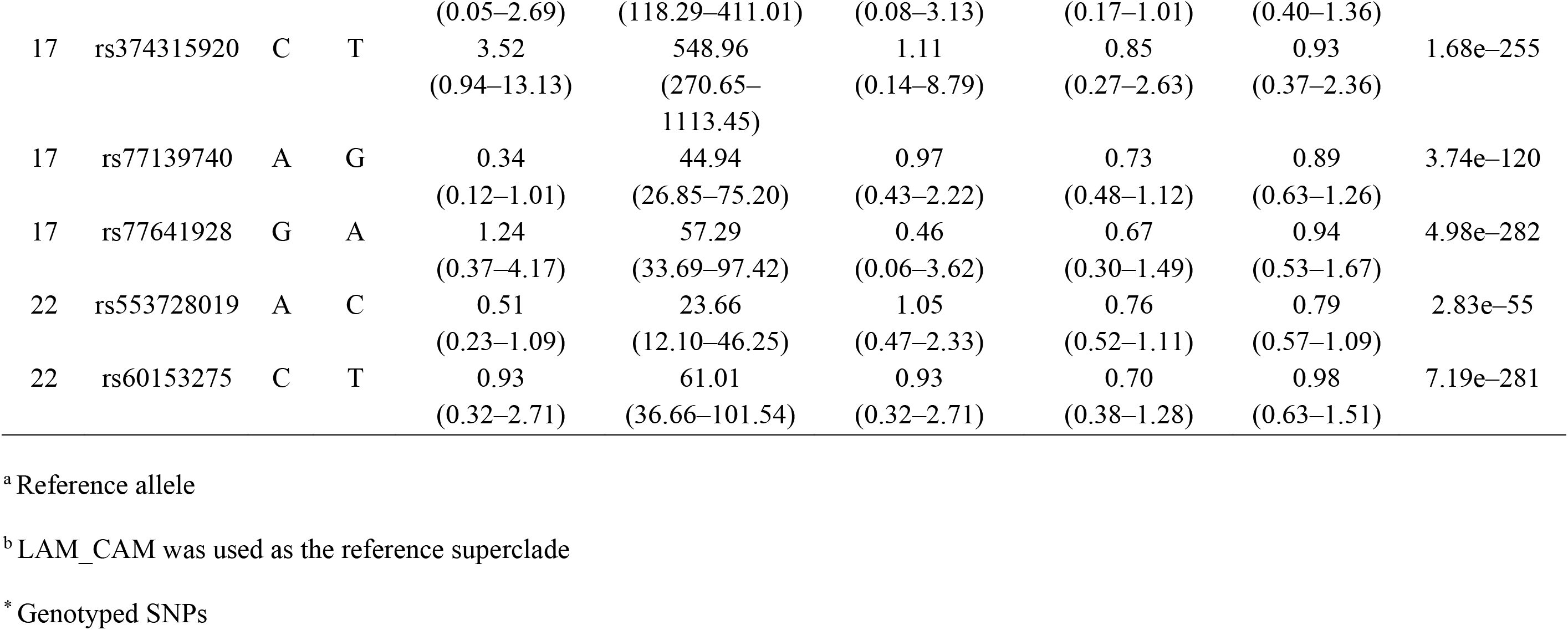
Top 32 SNPs identified by MLR to be associated with strains of different MTBC superclades in the Ghanaian cohort.

Several imputed SNPs were shown to dramatically increase the risk of being infected by a particular superclade. For example, the risk allele of the SNP rs577800201 (g.20476046C>T) was shown to increase the risk of being infected with the EAI_afri superclade by 93 times, compared to the baseline LAM_CAM superclade, and was annotated by the VEP to map to *ACSM2A*. The risk allele of the SNP rs374315920 (g.38496435C>T) located on chromosome 17, was also found to increase an individual’s risk of being infected with the EAI_afri superclade by more than 500 times, as compared to the LAMCAM reference superclade, and the MLR specific for this SNP was highly significant with an LRT p-value of 1.68 x 10^−255^. Using the VEP tool, this SNP was annotated to lie within the retinoic acid receptor alpha (RARA) gene. Since these SNPs were imputed, their associations should be considered tentative, because they may have been incorrectly imputed into this study dataset.

Further analysis revealed that of the 32 SNPs identified here, two, namely the intergenic variant rs529920 on chromosome 6, and the intron variant rs41472447 on chromosome 12, were directly genotyped. The risk allele of the SNP rs529920 (g.153835125A>G) was found to half the risk of being infected with a member of the BeijingCAS and EAI_afri superclades when compared to the LAM_CAM baseline superclade. Individuals with the risk allele for this SNP were also equally at risk of being infected with a member of the Ghana2, HaarlemX, or T_U superclades (OR: 1.04-1.24) (Table 5). Additionally, this SNP has an MAF of 0.4721 in the study cohort, which is similar to the MAF observed in African populations in the 1000G (MAF: 0.571) and in gnomAD genome (MAF:0.582). However, this SNP has no known gene consequence to date.

Similarly, the risk allele of the intron variant rs41472447 (g. 41708761A>G) was found to nearly triple the risk of infection with a member of the BeijingCAS superclade (OR: 2.56, C.I.: (1.48–4.41)) or the Ghana2 superclade (OR:2.94, C.I.: (1.71–5.08)). In contrast, the risk allele halved the risk of infection with the T_U superclade (OR:0.59, C.I.: 0.44-0.79) and had no effect on the risk for infection with members of the EAI_afri or HaarlemX superclades (Table 5). This variant had an MAF of 0.2461 in the study dataset which is similar to the MAF observed in African populations in the 1000G (MAF: 0.200) and in the gnomAD genome (MAF:0.165). Furthermore, rs41472447 maps to the *PDZRN4* gene in dbSNP, but has to date not been linked to susceptibility to tuberculosis.

## 3. Discussion

TB is a highly infectious disease affecting millions of people each year. The genetic susceptibility of the host to developing the disease has been extensively studied using linkage analysis, candidate gene studies, and GWAS. Furthermore, a number of selected genes have been investigated for their contribution to genetic susceptibility of the host to strains of different lineages of the MTBC. However, to date, no genome-wide analysis of genetic markers affecting susceptibility to strains of different MTBC lineages has been performed.

### 3.1. Imputation

While several reference panels exist to facilitate genotype imputation, most of these panels focus on representing populations of European ancestry, and little representation has been made for African populations. Therefore, the present study focused on evaluating the quality of imputation attainable for the five-way admixed South African population and the Ghanaian cohort using the 1000G, AGR, and CAAPA reference panels. The more admixed a population is, the greater the heterogeneity in its haplotype block structure. This genetic complexity requires large reference panels with suitable ancestry to facilitate accurate genotype imputation (Lin et al., n.d.). The five-way admixed South African population contains genetic contributions from Bantu-speaking Africans, Europeans, KhoeSan, and South- and East-Asians (Daya et al. 2013; De Wit et al. 2010). Imputation was previously performed for this population; however, it was done using the 1000G Phase 1 (The 1000 Genomes Project Consortium 2012) and the HapMap3 release 2 (The International HapMap 3 Consortium 2010) reference panels. The 1000G panel has been expanded substantially since whereas the HapMap3 reference panel, representing individuals mostly of European ancestry (Chimusa et al. 2014), has since been deprecated.

When evaluating the Ghanaian cohort, although imputation of the 1000G reference panel with the IH method performed the best, there was very little difference in the median quality scores for the different workflows seen for SNPs with an MAF of 10-50% (Fig. 5). For rare variants (MAF 0-5%) however, the IH method outperformed all others with a median quality score above 0.75, whereas both analyses with the MIS produced a median score below the cut-off of 0.45. Thus, for the Ghanaian cohort, all reference panels and methods tested could be considered viable options for imputing common variants with an MAF of 10-50% but should be considered carefully for variants with an MAF below 10%.

In contrast to the AGR which contains no individuals recruited from West-African countries, the CAAPA resource contains 88 individuals recruited from the West-African country of Nigeria (Mathias et al., 2016). Thus, it was unsurprising that the CAAPA resource performed well when imputing SNPs with a MAF above 10% (Fig. 5). From an MAF of 20-50%, the CAAPA resource performed similarly to the other three reference panels and may thus be considered suitable for imputing cohorts of West-African ancestry, such as the Ghanaian cohort used in this study.

### 3.2. Multi-phenotype GWAS

In this study, the MLR functionality within SNPTEST enabled the genome-wide investigation of genetic markers for association to a number of MTBC superclades. Although none of the SNPs passed the GWAS p-value cut-off, the most significant associations for the South African cohort imputed with the AGR reference panel were reported. This was likely due to the small sample size resulting in a subsequent reduction in statistical power. In contrast, 32 SNPs passed the GWAS cut-off for the Ghanaian cohort and may be considered as potential targets for further investigation of host-directed therapies suitable for individuals of West-African descent. None of the SNPs with an LRT p-value less than 0.0005 in either cohort were found in the other, demonstrating the population-specific association of SNPs with the strains of different MTBC superclades, which has been previously shown in the investigation of TLRs and their association with cases of TB in populations of different ethnicities (Schurz et al. 2015).

### 3.3. Potential drug targets

For the Ghanaian cohort, 32 SNPs with significant LRT p-values were identified as being associated with the MTBC superclades investigated (Table 5). Nine of the SNPs located on chromosome 12 mapped to the *PDZRN4* gene. For these nine SNPs, the risk allele increased the chances of individuals being infected with the BeijingCAS superclades 2.5 times, and in the region of three times for the Ghana2 superclade, while the risk allele halved the chances of being infected with the T_U superclade. Due to the low frequencies of the BeijingCAS and Ghana2 superclades observed for this cohort (Fig. 2D), it is possible that these odds ratios were inflated because of small sample sizes. Notably, the two SNPs which were directly genotyped, and not imputed, were found to be significantly associated with infection with particular MTBC superclades. This provided crucial evidence that despite the vast amount of imputed genotype data included in the MLR, the association analysis was able to detect two directly genotyped SNPs with potentially significant associations with the MTBC superclades.

Few studies have described the direct influence of *M. tb* infection on the expression of the *STARD4* and *RARA* genes, and no studies have investigated the outcomes of infection with strains belonging to different MTBC clades on these genes. The *STARD4* encodes the StAR-related lipid transfer protein which plays a crucial role in the transmembrane trafficking of lipids (such as cholesterol) - an important source of energy for *M. tb* (Soccio et al. 2002). Infection of macrophages with pathogens such as *M. tb* stimulates the process of lipid droplet formation (Daniel et al. 2011). It has been hypothesised that *M. tb* initiates this process in order to secure a reliable source of carbon to fuel bacterial growth (Brzostek et al. 2009). Additionally, the accumulation of cholesterol in the bacterial cell wall drastically reduces the permeability of the cell wall, subsequently reducing the penetrating capability of the anti-TB drug Rifampicin (Brzostek et al. 2009). However, a recent study has contradicted the notion that lipid droplet formation is a bacteria-driven process. Instead, it was proposed that the formation of lipids is an immune system-activated process, and does not occur as a result of direct stimulation by *M. tb*, but rather via the IFN-ƴ, H1F-alpha-dependent pathway of the host immune system (Knight et al. 2018).

All-trans retinoic acid (*RARA*), the active form of Vitamin A, plays an essential role in the normal functioning of the adaptive and innate immune systems. The oral administration of retinoic acid to rats resulted in inhibition of the *M. tb* growth, following *in vitro* infection (Yamada et al. 2007), thus making this gene a potential target for anti-TB therapies. The results of this study have highlighted several SNPs which possibly significantly increased the risk of individuals with Ghanaian ethnicity to being infected with the endemic TB strain of *M. africanum*. Given the burden of disease, and the dominance of *M. africanum* strains in Ghana, it may be a worthwhile exploring the functional effect of these SNPs on the biological processes described.

### 3.4. Recommended improvements for future multi-phenotype GWAS

Several limitations may have affected this study. Obtaining a suitable sample size is a problem inherent in GWA studies making use of logistic regression modelling. Furthermore, the many phenotypes being analysed in this study, demanded a sufficient number of cases for each class. The frequency of each MTBC clade however, is dependent not only on the host, but is also affected by the virulence of the bacterium. Thus, with all these considered, the sample sizes included in the MLR should be sufficient for inclusion in the analysis but will likely also reflect the distribution in the population. Another limitation of the study is that at the time of analysis, the AGR reference panel was not publicly available for download to a local machine. Thus, its use in this study could only be facilitated via the SIS, a freely accessible online imputation server. Through this, we were able to obtain high-quality imputed data for the South African dataset, but it was necessary to be mindful when drawing comparisons as the other workflows made use of different imputation software.

With the current trajectory of the TB epidemic, novel methods are needed to augment current therapies for TB and combat the disease. This study provides the groundwork for future GWAS wishing to investigate the relationship between the host and the many members of the MTBC causing disease. Furthermore, the SNPs identified in this study may be evaluated in functional studies to assess their viability as targets for host-directed therapies. This study would not have been possible were it not for the collection of paired samples of blood and sputum from study participants. Thus, future studies of this kind will require that both samples be collected from participants in order to perform this association analysis. Although it was necessary to exclude low frequency MTBC cases at the superclade level to prevent the reduction in statistical power of the association test, this may have brought in a weakness in interpreting the odds ratios derived from the model. Thus, the odds ratios derived may only be interpreted at the superclade level and does not provide further granularity to association with specific clades. Therefore, future studies employing this method should consider excluding low-frequency clades before clustering as well as Bayesian analyses that allow inclusion of prior probability distribution for strain prevalence. Additionally, incorporation of more diverse reference panels, such as the AGR, and new algorithms for imputation could improve association results.

## 4. Materials and methods

### 4.1. Study design

To perform genome-wide association analyses between human host genotypes and the infecting member of the MTBC, host genotypes with paired MTBC isolate information were sourced for two geographically distinct cohorts. Paired host genotype and isolate information was available for a South African cohort, and a Ghanaian cohort. As these two cohorts are geographically distinct, and possess vastly different admixture profiles, we did not aim to replicate our findings within these two cohorts.

The South African cohort consisted of study participants recruited in the Western Cape Province of South Africa during the period of January 1993 through December 2004. Participants were recruited from suburbs where the TB incidence was high (28.9 % in 2005) and the prevalence of HIV was a low 2% (Kritzinger et al., 2009; Shisana et al., 2012). All study participants in this cohort self-identified as belonging to a five-way admixed South African population, were HIV-negative, and provided written informed consent. Blood samples were collected for SNP genotyping of the host and sputum samples were collected for bacterial culture on Loewenstein-Jensen (L-J) media. For the Ghanaian cohort, participants were enrolled between September 2001 and July 2004 at Korle Bu Teaching Hospital in Accra, Komfo Anokye Teaching Hospital in Kumasi, and at 15 additional hospitals and polyclinics in Accra and Kumasi, as well as regional district hospitals. All cases were HIV-negative and confirmed to have pulmonary TB by sputum microscopy, performing solid mycobacterial cultures using L-J media and also by two independent radiologists (Thye et al., 2012, 2010). All subsequent research was conducted in accordance with the principles expressed in the Declaration of Helsinki (WHO, 2001).

### 4.2. Host SNP genotyping

#### 4.2.1. South Africa

SNP genotyping was performed using DNA extracted from blood samples. All participants in the South African cohort were genotyped using the GeneChip Human Mapping 500K SNP array which contains 500 000 SNP markers (Affymetrix, California, United States), while a subset of this cohort was also genotyped using the Infinium Multi-Ethnic Genotyping Array (MEGA), which is comprised of 1.7 million SNP markers (Illumina, California, United States). Genotype-calling was performed using the Affymetrix Power Tools pipeline (V1.10.0) as previously described (De Wit et al., 2010; Schurz et al., 2019a). Genotype datum was made available in PLINK format (Purcell and Chang, n.d.; Chang et al. 2015). Following standard genotyping quality control (QC), ancestry proportions for the South African cohort on both the Affymetrix and MEGA arrays were estimated (Daya et al. 2013, Schurz et al. 2018) using the unsupervised algorithm implemented in ADMIXTURE (Alexander et al. 2009).

#### 4.2.2. Ghana

DNA extracted from blood samples was genotyped using the Affymetrix SNP 6.0 array at the Affymetrix Services Laboratory in California, and at ATLAS Biolabs GmbH in Berlin. Genotype datum was made available in PLINK format, genotypes were called using the Birdseed version 2 algorithm and ancestry proportions in the form of principal components were derived using the Eigenstrat software (Thye et al., 2012).

### 4.3. Bacterial SNP genotyping

For the South African cohort, MTBC isolates were genotyped using spoligotyping and IS*6110* Restriction Fragment Length Polymorphism (RFLP) methods as previously described (van der Spuy et al. 2009). For the Ghanaian cohort, MTBC isolates extracted from sputum samples were cultured on L-J media at the Kumasi Centre for Collaborative Research (Owusu-Dabo et al., 2006) and strains were identified using IS*6110* RFLP and spoligotyping (Supply et al., 2006). All MTBC isolate information were captured on manually-curated infection databases for archiving and made available for this study.

### 4.4. Defining MTBC clades and superclades

The term “superclades” was used to describe the grouping of clades using a SNP-based phylogenetic tree of the MTBC. Where a common progenitor was shared, MTBC clades were grouped into superclades near a point of divergence (Fig. 1) (Dippenaar, 2014). This was performed to reduce the number of clades of low frequency, as low frequency groups are known to induce an unfavourable collinearity effect on logistic regression models (Bergtold et al., 2011). Additionally, clustering of clades into superclades mitigated the class imbalance which would negatively affect the statistical models. Clades not amalgamated with other clades were also referred to as “superclades” after clustering and those not represented on the phylogenetic tree, were kept as distinct superclades and not clustered with any of the members on the existing tree, except when a suitable reference phylogeny was found. After clustering, superclades with a frequency less than ten in the study dataset were excluded from subsequent analyses.

### 4.5. Genotype data quality control, haplotype phasing, and genotype imputation

Quality control, haplotype phasing and genotype imputation were performed using the methods described in Schurz *et al*. (2019). Briefly, genotype data were iteratively filtered for 2% SNP genotype missingness, 10% sample missingness and 5% SNP minor allele frequency (MAF), until no additional samples or variants were removed. Additionally, variants lacking chromosome or base pair information were updated using the 1000 Genomes Phase 3 (1000G) reference panel and the dbSNP (Sherry et al., 2001) database. Imputation processes are strengthened by sample size. Thus, all available samples, regardless of relatedness or whether the sample had matching MTBC information, were included in the imputation. Related individuals were noted, but not removed in order to maximise the number of haplotypes available for the imputation process. The genotype QC procedure concluded with a sex concordance check as well as strand-alignment to the 1000G reference panel [human genome build 37 (Sudmant et al., 2015)] using the Genotype Harmonizer (version 1.4.15) tool (Deelen et al., 2014).

Following the initial QC, haplotype phasing was performed using the ShapeITv2 (Delaneau et al., 2008) software set at default parameters. Although some studies have reported that pre-phasing reduces imputation accuracy (Roshyara et al. 2016), it is known to significantly speed up the computationally intensive process of genotype imputation (Kanterakis et al. 2015; B. Howie et al. 2012). To maximise the number of variants tested in the association analysis, the cleaned genotype data were imputed using five protocols - and three different reference panels - to determine which panel best served the given dataset in imputing missing variants, as detailed in Schurz *et al*. (2019).

The In-House (IH) protocol made use of the 1000G reference panel with the IMPUTE2 imputation software (Howie et al., 2009). The Sanger Imputation Server (SIS) (McCarthy et al., 2016) makes use of the Positional Burrows-Wheeler Transformation (PBWT) algorithm (Durbin, 2014), with two options for the reference panel: the African Genome Resource (AGR) and the 1000G. Lastly, the Michigan Imputation Server (MIS) (Das et al., 2016) makes use of the Minimac3 algorithm (Howie et al., 2009) and provides access to imputation using the 1000G and the Consortium on Asthma among African-ancestry Populations in the Americas (CAAPA) (Mathias et al., 2016) reference panels. Of note, both the CAAPA and AGR reference panels were not publicly accessible for download at the time of this study and thus could only be accessed using these two online imputation server platforms. The key differences between the three protocols using the 1000G reference panel was the imputation software used, as well as additional strict QC filters imposed on the study dataset by the MIS and SIS methods.

### 4.6. Selection of high-quality imputed genotype data

The quality control procedure for the imputed data was implemented as described in Schurz *et al*. (2019). Briefly, following five imputation protocols, imputed data were filtered using a genotype calling threshold of 0.7, and the internal quality metric produced by the imputation process (Schurz et al., 2019b). SNPs with an INFO or R-squared (Rsq) value greater than 0.45 were prioritised for the association analysis and filtered iteratively for a maximum of 2% SNP genotype missingness, 10% sample missingness, and 5% SNP MAF using PLINK. Related individuals identified prior to imputation were removed followed by a second round of iterative filters for SNP- and sample missingness and MAF (Schurz et al., 2019b). Post-imputation QC concluded with extracting MTBC clade-matched samples from the remaining samples which had passed all QC filters.

### 4.7. Covariable data

All available covariables were obtained including sex, and age at time of active TB and subsequent recruitment into the study. To correct for differences in ethnicity amongst participants, either ancestry proportions or principal components were calculated and included as covariables.

### 4.8. Multi-phenotype GWAS

For the association analysis, a multinomial logistic regression (MLR) analysis using an additive genetic model was performed using SNPTEST v2.5.2 (Marchini, 2010). Two discrete variables, namely sex and superclade, as well as continuous variables, namely age at TB onset and ancestry proportions or principal components were included in the analysis. The phenotype tested was specified as the MTBC superclade. Thus, the MLR model specified was the occurrence of the MTBC superclade as a function of the baseline covariables given, as well as the host genotypes supplied.

SNPTEST is unable to include covariables when the variance in the values provided is “too small” as indicated (Marchini, n.d.). At the time of this study, SNPTEST was still under development, and it had not yet been established, or recorded in the software manual, to what degree of variance covariable data would not be accepted for inclusion in the logistic regression. For developing the method, it was established through trial and error that if the variance was below 0.001, these covariables could not be included.

The standard genome-wide significance cut-off of alpha = 5×10^-8^ was used when reporting significance of SNPs (Pe’er et al., 2008; The International HapMap Consortium, 2005). Odds ratios (OR) for the multiple phenotypes tested were calculated against a baseline phenotype by setting the odds of that phenotype occurring, given the genotype, to 1. For this study, the baseline phenotype was specified as the dominant superclade in the cohort, or a common superclade of intermediate frequency if more than one cohort was being studied. Thus, the LAM- and LAM_CAM superclades were used as the baseline phenotype for the association analyses of the South African, and Ghanaian cohorts, respectively.

SNPs with a Likelihood Ratio Threshold (LRT) p-value of less than 5×10^-4^ were selected and analysed in R (R Core Team, 2017) and odds ratios were calculated from the beta values generated by SNPTEST. SNPs with a standard error greater than 1.5 for their odds ratios were excluded and SNPs with an LRT p-value less than 1×10^-6^ were prioritised for further investigation. These thresholds were chosen pragmatically to facilitate the completion of method development. Finally, the Variant Effect Predictor (VEP) Tool (McLaren et al. 2016) was used to retrieve gene annotations for the SNPs of interest.

## Data Availability

Summary statistics for the South African cohorts data can be made available to researchers who meet the criteria for access to confidential data after application to the Health Research Ethics Committee of Stellenbosch University. Requests may be sent to: Prof Craig Kinnear, E mail

## Acknowledgements

The authors would like to acknowledge and thank the study participants for their contribution and participation. This research was partially funded by the South African government through the South African Medical Research Council. This work was also supported by the National Research Foundation of South Africa (grant number 93460) to E.H. and by a Strategic Health Innovation Partnership grant from the South African Medical Research Council and Department of Science and Innovation/South African Tuberculosis Bioinformatics Initiative to G.T. The content is solely the responsibility of the authors and does not necessarily represent the official views of the South African Medical Research Council.

## Ethics approval and consent to participate

For the South African cohort, blood and sputum samples were collected from study participants as approved by the Health Research Ethics Committee of Stellenbosch University (Project numbers: S17/01/013 and 95/072). For the Ghanaian cohort, ethics for the study protocol was granted by the Committee on Human Research, Publications and Ethics, School of Medical Sciences, Kwame Nkrumah University of Science and Technology, Kumasi, Ghana, and the Ethics Committee of the Ghana Health Service, Accra, Ghana (Thye et al., 2012). Venous blood samples were taken only after a detailed explanation of the aims of the study, and consent was obtained of individuals enrolled or their parents/guardians by signature or by thumbprint in case of illiteracy.

## Availability of data and materials

Summary statistics for the South African cohort’s data can be made available to researchers who meet the criteria for access to confidential data after application to the Health Research Ethics Committee of Stellenbosch University. Requests may be sent to: Prof Craig Kinnear.

## Author contributions

**Stephanie J. Müller**: Conceptualization, Methodology, Software, Investigation, Formal analysis, Writing - Original Draft, Visualization

**Haiko Schurz:** Software, Validation, Writing - Review & Editing

**Gerard Tromp:** Conceptualization, Supervision, Visualization, Writing - Review & Editing

**Gian D. van der Spuy:** Conceptualization, Supervision, Data Curation, Resources, Writing - Review & Editing

**Eileen G. Hoal:** Conceptualization, Supervision, Writing - Review & Editing

**Paul D. van Helden:** Conceptualization, Writing - Review & Editing

**Ellis Owusu-Dabo:** Data Curation, Writing - Review & Editing

**Christian G. Meyer:** Data Curation, Writing - Review & Editing

**Thorsten Thye:** Data Curation, Resources, Writing - Review & Editing

**Stefan Niemann:** Data Curation, Writing - Review & Editing

**Robin M. Warren:** Data Curation, Resources, Writing - Review & Editing

**Elizabeth Streicher:** Data Curation, Writing - Review & Editing

**Marlo Möller:** Conceptualization, Supervision, Writing - Review & Editing

**Craig Kinnear:** Conceptualization, Supervision, Writing - Review & Editing

## References

Alexander, D.H., Novembre, J., Lange, K., 2009. Fast Model-Based Estimation of Ancestry in Unrelated Individuals. Genome Res. 1655–1664. https://doi.org/10.1101/gr.094052.109.vidual

Asante-Poku, A., Yeboah-Manu, D., Otchere, I.D., Aboagye, S.Y., Stucki, D., Hattendorf, J., Borrell, S., Feldmann, J., Danso, E., Gagneux, S., 2015. Mycobacterium africanum Is Associated with Patient Ethnicity in Ghana. PLoS Negl. Trop. Dis. 9, e3370. https://doi.org/10.1371/journal.pntd.0003370

Bellamy, R., 1998. Genetic susceptibility to tuberculosis in human populations. Thorax 53, 588–593.

Bergtold, J.S., Yeager, E.A., Featherstone, A., 2011. Sample Size and Robustness of Inferences from Logistic Regression in the Presence of Nonlinearity and Multicollinearity. Presented at the Agricultural & Applied Economics Association’s 2011 AAEA & NAREA Joint Annual Meeting, Pittsburgh, Pennsylvania.

Brites, D., Gagneux, S., 2015. Co-evolution of Mycobacterium tuberculosis and Homo sapiens. Immunol. Rev. 264, 6–24. https://doi.org/10.1111/imr.12264

Caws, M., Thwaites, G., Dunstan, S., Hawn, T.R., Thi Ngoc Lan, N., Thuong, N.T.T., Stepniewska, K., Huyen, M.N.T., Bang, N.D., Huu Loc, T., Gagneux, S., van Soolingen, D., Kremer, K., van der Sande, M., Small, P., Thi Hoang Anh, P., Chinh, N.T., Thi Quy, H., Thi Hong Duyen, N., Quang Tho, D., Hieu, N.T., Torok, E., Hien, T.T., Dung, N.H., Thi Quynh Nhu, N., Duy, P.M., van Vinh Chau, N., Farrar, J., 2008. The Influence of Host and Bacterial Genotype on the Development of Disseminated Disease with Mycobacterium tuberculosis. PLoS Pathog. 4. https://doi.org/10.1371/journal.ppat.1000034

Chang, C.C., Chow, C.C., Tellier, L.C., Vattikuti, S., Purcell, S.M., Lee, J.J., 2015. Second-generation PLINK: rising to the challenge of larger and richer datasets. GigaScience 4. https://doi.org/10.1186/s13742-015-0047-8

Cheng, M.P., Abou Chakra, C.N., Yansouni, C.P., Cnossen, S., Shrier, I., Menzies, D., Greenaway, C., 2016. Risk of Active Tuberculosis in Patients with Cancer: A Systematic Review and Meta-Analysis. Clin. Infect. Dis. ciw838. https://doi.org/10.1093/cid/ciw838

Chimusa, E.R., Zaitlen, N., Daya, M., Möller, M., Helden, P.D. van Nicola, J.M., Price, A.L., Hoal, E.G., 2014. Genome-wide association study of ancestry-specific TB risk in the South African coloured population. Hum. Mol. Genet. 23, 796–809. https://doi.org/10.1093/hmg/ddt462

Das, S., Forer, L., Schönherr, S., Sidore, C., Locke, A.E., Kwong, A., Vrieze, S.I., Chew, E.Y., Levy, S., McGue, M., Schlessinger, D., Stambolian, D., Loh, P.-R., Iacono, W.G., Swaroop, A., Scott, L.J., Cucca, F., Kronenberg, F., Boehnke, M., Abecasis, G.R., Fuchsberger, C., 2016. Next-generation genotype imputation service and methods. Nat. Genet. 48, 1284–1287. https://doi.org/10.1038/ng.3656

Daya, M., Van Der Merwe, L., Galal, U., Möller, M., Salie, M., Chimusa, E.R., Galanter, J.M., Van Helden, P.D., Henn, B.M., Gignoux, C.R., Hoal, E., 2013. A panel of ancestry informative markers for the complex five-way admixed South African Coloured population. PLoS ONE 8, 12–12. https://doi.org/10.1371/journal.pone.0082224

De Wit, E., Delport, W., Rugamika, C.E., Meintjes, A., Moller, M., Van Helden, P.D., Seoighe, C., Hoal, E.G., 2010. Genome-wide analysis of the structure of the South African Coloured Population in the Western Cape. Hum. Genet. 128, 145–153. https://doi.org/10.1007/s00439-010-0836-1

Deelen, P., Bonder, M.J., van der Velde, K.J., Westra, H.-J., Winder, E., Hendriksen, D., Franke, L., Swertz, M.A., 2014. Genotype harmonizer: automatic strand alignment and format conversion for genotype data integration. BMC Res. Notes 7, 901.

Delaneau, O., Coulonges, C., Zagury, J.-F., 2008. Shape-IT: new rapid and accurate algorithm for haplotype inference. BMC Bioinformatics 9, 540. https://doi.org/10.1186/1471-2105-9-540

Dippenaar, A., 2014. A phylogenomic-and proteomic investigation into the evolution and biological characteristics of the members of the group 2 Latin-American Mediterranean (LAM) genotype of Mycobacterium tuberculosis (PhD Thesis). Stellenbosch: Stellenbosch University.

Durbin, R., 2014. Efficient haplotype matching and storage using the positional Burrows–Wheeler transform (PBWT). Bioinformatics 30, 1266–1272. https://doi.org/10.1093/bioinformatics/btu014

Gagneux, S., 2012. Host-pathogen coevolution in human tuberculosis. Philos. Trans. R. Soc. Lond. B. Biol. Sci. 367, 850–9. https://doi.org/10.1098/rstb.2011.0316

Gagneux, S., Small, P.M., 2007. Global phylogeography of Mycobacterium tuberculosis and implications for tuberculosis product development. Lancet Infect. Dis. 7, 328–337. https://doi.org/10.1016/S1473-3099(07)70108-1

Herb, F., Thye, T., Niemann, S., Browne, E.N.L., Chinbuah, M.A., Gyapong, J., Osei, I., Owusu-Dabo, E., Werz, O., Rusch-Gerdes, S., Horstmann, R.D., Meyer, C.G., 2007. ALOX5 variants associated with susceptibility to human pulmonary tuberculosis. Hum. Mol. Genet. 17, 1052–1060. https://doi.org/10.1093/hmg/ddm378

Hirschhorn, J.N., Daly, M.J., 2005. Genome-wide association studies for common diseases and complex traits. Nat. Rev. Genet. 6, 95–108. https://doi.org/10.1038/nrg1521

Hoal, E.G., Dippenaar, A., Kinnear, C., van Helden, P.D., Möller, M., 2017. The arms race between man and Mycobacterium tuberculosis: Time to regroup. Infect. Genet. Evol. https://doi.org/10.1016/j.meegid.2017.08.021

Hong, E.P., Go, M.J., Kim, H.-L., Park, J.W., 2017. Risk prediction of pulmonary tuberculosis using genetic and conventional risk factors in adult Korean population. PLOS ONE 12, e0174642. https://doi.org/10.1371/journal.pone.0174642

Howie, B.N., Donnelly, P., Marchini, J., 2009. A Flexible and Accurate Genotype Imputation Method for the Next Generation of Genome-Wide Association Studies. PLOS Genet. 5, e1000529. https://doi.org/10.1371/journal.pgen.1000529

Kinnear, C., Hoal, E.G., Schurz, H., Van Helden, P.D., Moller, M., 2017. The role of human host genetics in tuberculosis resistance. Expert Rev. Respir. Med. 11, 721–737. https://doi.org/10.1080/17476348.2017.1354700

Kritzinger, F.E., den Boon, S., Verver, S., Enarson, D.A., Lombard, C.J., Borgdorff, M.W., Gie, R.P., Beyers, N., 2009. No decrease in annual risk of tuberculosis infection in endemic area in Cape Town, South Africa. Trop. Med. Int. Health 14, 136–142. https://doi.org/10.1111/j.1365-3156.2008.02213.x

Lim, C.H., Chen, H.-H., Chen, Y.-H., Chen, D.-Y., Huang, W.-N., Tsai, J.-J., Hsieh, T.-Y., Hsieh, C.-W., Hung, W.-T., Lin, C.-T., Lai, K.-L., Tang, K.-T., Tseng, C.-W., Chen, Y.-M., 2017. The risk of tuberculosis disease in rheumatoid arthritis patients on biologics and targeted therapy: A 15-year real world experience in Taiwan. PLOS ONE 12, e0178035. https://doi.org/10.1371/journal.pone.0178035

Lin, M., Caberto, C., Wan, P., Li, Y., Lum-Jones, A., Tiirikainen, M., Pooler, L., Nakamura, B., Sheng, X., Porcel, J., Lim, U., Setiawan, V.W., Le Marchand, L., Wilkens, L.R., Haiman, C.A., Cheng, I., Chiang, C.W.K., n.d. Population-specific reference panels are crucial for genetic analyses: an example of the CREBRF locus in Native Hawaiians. Hum. Mol. Genet. https://doi.org/10.1093/hmg/ddaa083

Marchini, J., 2010. SNPTEST v2 Technical Details 10.

Marchini, J., n.d. SNPTest.

Mathias, R.A., Taub, M.A., Gignoux, C.R., Fu, W., Musharoff, S., O’Connor, T.D., Vergara, C., Torgerson, D.G., Pino-Yanes, M., Shringarpure, S.S., Huang, L., Rafaels, N., Boorgula, M.P., Johnston, H.R., Ortega, V.E., Levin, A.M., Song, W., Torres, R., Padhukasahasram, B., Eng, C., Mejia-Mejia, D.-A., Ferguson, T., Qin, Z.S., Scott, A.F., Yazdanbakhsh, M., Wilson, J.G., Marrugo, J., Lange, L.A., Kumar, R., Avila, P.C., Williams, L.K., Watson, H., Ware, L.B., Olopade, C., Olopade, O., Oliveira, R., Ober, C., Nicolae, D.L., Meyers, D., Mayorga, A., Knight-Madden, J., Hartert, T., Hansel, N.N., Foreman, M.G., Ford, J.G., Faruque, M.U., Dunston, G.M., Caraballo, L., Burchard, E.G., Bleecker, E., Araujo, M.I., Herrera-Paz, E.F., Gietzen, K., Grus, W.E., Bamshad, M., Bustamante, C.D., Kenny, E.E., Hernandez, R.D., Beaty, T.H., Ruczinski, I., Akey, J., Barnes, K.C., 2016. A continuum of admixture in the Western Hemisphere revealed by the African Diaspora genome. Nat. Commun. 7. https://doi.org/10.1038/ncomms12522

McCarthy, S., Das, S., Kretzschmar, W., Delaneau, O., Wood, A.R., Teumer, A., Kang, H.M., Fuchsberger, C., Danecek, P., Sharp, K., Luo, Y., Sidore, C., Kwong, A., Timpson, N., Koskinen, S., Vrieze, S., Scott, L.J., Zhang, H., Mahajan, A., Veldink, J., Peters, U., Pato, C., van Duijn, C.M., Gillies, C.E., Gandin, I., Mezzavilla, M., Gilly, A., Cocca, M., Traglia, M., Angius, A., Barrett, J.C., Boomsma, D., Branham, K., Breen, G., Brummett, C.M., Busonero, F., Campbell, H., Chan, A., Chen, S., Chew, E., Collins, F.S., Corbin, L.J., Smith, G.D., Dedoussis, G., Dorr, M., Farmaki, A.-E., Ferrucci, L., Forer, L., Fraser, R.M., Gabriel, S., Levy, S., Groop, L., Harrison, T., Hattersley, A., Holmen, O.L., Hveem, K., Kretzler, M., Lee, J.C., McGue, M., Meitinger, T., Melzer, D., Min, J.L., Mohlke, K.L., Vincent, J.B., Nauck, M., Nickerson, D., Palotie, A., Pato, M., Pirastu, N., McInnis, M., Richards, J.B., Sala, C., Salomaa, V., Schlessinger, D., Schoenherr, S., Slagboom, P.E., Small, K., Spector, T., Stambolian, D., Tuke, M., Tuomilehto, J., Van den Berg, L.H., Van Rheenen, W., Volker, U., Wijmenga, C., Toniolo, D., Zeggini, E., Gasparini, P., Sampson, M.G., Wilson, J.F., Frayling, T., de Bakker, P.I.W., Swertz, M.A., McCarroll, S., Kooperberg, C., Dekker, A., Altshuler, D., Willer, C., Iacono, W., Ripatti, S., Soranzo, N., Walter, K., Swaroop, A., Cucca, F., Anderson, C.A., Myers, R.M., Boehnke, M., McCarthy, M.I., Durbin, R., Abecasis, G., Marchini, J., the Haplotype Reference Consortium, 2016. A reference panel of 64,976 haplotypes for genotype imputation. Nat. Genet. 48, 1279–1283. https://doi.org/10.1038/ng.3643

McHenry, M.L., Bartlett, J., Igo, R.P., Wampande, E., Benchek, P., Mayanja-Kizza, H., Fluegge, K., Hall, N.B., Gagneux, S., Tishkoff, S.A., Wejse, C., Sirugo, G., Boom, W.H., Joloba, M., Williams, S.M., Stein, C.M., 2019. Interaction between host genes and M. tuberculosis lineage can affect tuberculosis severity: evidence for coevolution. bioRxiv 769448. https://doi.org/10.1101/769448

Möller, M., Hoal, E.G., 2010. Current findings, challenges and novel approaches in human genetic susceptibility to tuberculosis. Tuberculosis 90, 71–83. https://doi.org/10.1016/j.tube.2010.02.002

Oki, N.O., Motsinger-Reif, A.A., Antas, P.R., Levy, S., Holland, S.M., Sterling, T.R., 2011. Novel human genetic variants associated with extrapulmonary tuberculosis: a pilot genome wide association study. BMC Res. Notes 4, 28. https://doi.org/10.1186/1756-0500-4-28

Omae, Y., Toyo-oka, L., Yanai, H., Nedsuwan, S., Wattanapokayakit, S., Satproedprai, N., Smittipat, N., Palittapongarnpim, P., Sawanpanyalert, P., Inunchot, W., Pasomsub, E., Wichukchinda, N., Mushiroda, T., Kubo, M., Tokunaga, K., Mahasirimongkol, S., 2017. Pathogen lineage-based genome-wide association study identified CD53 as susceptible locus in tuberculosis. J. Hum. Genet. https://doi.org/10.1038/jhg.2017.82

Owusu-Dabo, E., Adjei, O., Meyer, C.G., Horstmann, R.D., Enimil, A., Kruppa, T.F., Bonsu, F., Browne, E.N.L., Chinbuah, M.A., Osei, I., Gyapong, J., Berberich, C., Kubica, T., Niemann, S., Ruesch-Gerdes, S., 2006. Mycobacterium tuberculosis Drug Resistance, Ghana. Emerg. Infect. Dis. 12, 1170–1172. https://doi.org/10.3201/eid1207.051028

Pastinen, T., Liitsola, K., Niini, P., Salminen, M., SyväNen, A.-C., 1998. Contribution of the CCR5 and MBL Genes to Susceptibility to HIV Type 1 Infection in the Finnish Population. AIDS Res. Hum. Retroviruses 14, 695–698. https://doi.org/10.1089/aid.1998.14.695

Pe’er, I., Yelensky, R., Altshuler, D., Daly, M.J., 2008. Estimation of the multiple testing burden for genomewide association studies of nearly all common variants. Genet. Epidemiol. 32, 381–385. https://doi.org/10.1002/gepi.20303

Png, E., Alisjahbana, B., Sahiratmadja, E., Marzuki, S., Nelwan, R., Balabanova, Y., Nikolayevskyy, V., Drobniewski, F., Nejentsev, S., Adnan, I., van de Vosse, E., Hibberd, M.L., van Crevel, R., Ottenhoff, T.H., Seielstad, M., 2012. A genome wide association study of pulmonary tuberculosis susceptibility in Indonesians. BMC Med. Genet. 13. https://doi.org/10.1186/1471-2350-13-5

Pratiwi, R.D., 2016. Socio-economic and environmenys risk factors of Tuberculosis in Wonosobo, Central Java, Indonesia. Graduate Studies in Public Health, Graduate Program, Sebelas Maret University Jl. Ir Sutami 36A, Surakarta 57126. Telp/Fax: (0271) 632 450 ext.208 First website:http//:s2ikm.pasca.uns.ac.id Second website: www.theicph.com., p. 89. https://doi.org/10.26911/theicph.2016.027

Purcell, S., Chang, C., n.d. PLINK 1.9.

R Core Team, 2017. R: A language and environment for statistical computing. R Foundation for Statistical Computing. Vienna, Austria.

Rockett, K.A., Clarke, G.M., Fitzpatrick, K., Hubbart, C., Jeffreys, A.E., Rowlands, K., Craik, R., Jallow, M., Conway, D.J., Bojang, K.A., Pinder, M., Usen, S., Sisay-Joof, F., Sirugo, G., Toure, O., Thera, M.A., Konate, S., Sissoko, S., Niangaly, A., Poudiougou, B., Mangano, V.D., Bougouma, E.C., Sirima, S.B., Modiano, D., Amenga-Etego, L.N., Ghansah, A., Koram, K.A., Wilson, M.D., Enimil, A., Evans, J., Amodu, O., Olaniyan, S., Apinjoh, T., Mugri, R., Ndi, A., Ndila, C.M., Uyoga, S., Macharia, A., Peshu, N., Williams, T.N., Manjurano, A., Riley, E., Drakeley, C., Reyburn, H., Nyirongo, V., Kachala, D., Molyneux, M., Dunstan, S.J., Phu, N.H., Ngoc Quyen, N.T., Thai, C.Q., Hien, T.T., Manning, L., Laman, M., Siba, P., Karunajeewa, H., Allen, S., Allen, A., Davis, T.M.E., Michon, P., Mueller, I., Green, A., Molloy, S., Johnson, K.J., Kerasidou, A., Cornelius, V., Hart, L., Vanderwal, A., SanJoaquin, M., Band, G., Le, S.Q., Pirinen, M., Sepúlveda, N., Spencer, C.C.A., Clark, T.G., Agbenyega, T., Achidi, E., Doumbo, O., Farrar, J., Marsh, K., Taylor, T., Kwiatkowski, D.P., 2014. Reappraisal of known malaria resistance loci in a large multi-centre study. Nat. Genet. 46, 1197–1204. https://doi.org/10.1038/ng.3107

Salie, M., Van Der Merwe, L., Moller, M., Daya, M., Van Der Spuy, G.D., Van Helden, P.D., Martin, M.P., Gao, X.J., Warren, R.M., Carrington, M., Hoal, E.G., 2014. Associations between human leukocyte antigen class i variants and the mycobacterium tuberculosis subtypes causing disease. J. Infect. Dis. 209, 216–223. https://doi.org/10.1093/infdis/jit443

Schurz, H., Kinnear, C.J., Gignoux, C., Wojcik, G., van Helden, P.D., Tromp, G., Henn, B., Hoal, E.G., Möller, M., 2019a. A Sex-Stratified Genome-Wide Association Study of Tuberculosis Using a Multi-Ethnic Genotyping Array. Front. Genet. 9. https://doi.org/10.3389/fgene.2018.00678

Schurz, H., Müller, S.J., van Helden, P.D., Tromp, G., Hoal, E.G., Kinnear, C.J., Möller, M., 2019b. Evaluating the Accuracy of Imputation Methods in a Five-Way Admixed Population. Front. Genet. 10. https://doi.org/10.3389/fgene.2019.00034

Seddon, J.A., Hesseling, A.C., Godfrey-Faussett, P., Fielding, K., Schaaf, H.S., 2013. Risk factors for infection and disease in child contacts of multidrug-resistant tuberculosis: a cross-sectional study. BMC Infect. Dis. 13. https://doi.org/10.1186/1471-2334-13-392

Sherry, S.T., Ward, M.-H., Kholodov, M., Baker, J., Phan, L., Smigielski, E.M., Sirotkin, K., 2001. dbSNP: the NCBI database of genetic variation. Nucleic Acids Res. 29, 308–311.

Shisana, O., Rhele, T., Simbayi, L.C., Zuma, K., Jooste, S., Zungu, N., Labadarios, D., Onoya, D., 2012. South African National HIV Prevalence, Incidence and Behaviour Survey, 2012.

Søborg, C., Madsen, H.O., Andersen, Å.B., Lillebaek, T., Kok-Jensen, A., Garred, P., 2003. Mannose-Binding Lectin Polymorphisms in Clinical Tuberculosis. J. Infect. Dis. 188, 777–782. https://doi.org/10.1086/377183

Stucki, D., Brites, D., Jeljeli, L., Coscolla, M., Liu, Q., Trauner, A., Fenner, L., Rutaihwa, L., Borrell, S., Luo, T., Gao, Q., Kato-Maeda, M., Ballif, M., Egger, M., Macedo, R., Mardassi, H., Moreno, M., Vilanova, G.T., Fyfe, J., Globan, M., Thomas, J., Jamieson, F., Guthrie, J.L., Asante-Poku, A., Yeboah-Manu, D., Wampande, E., Ssengooba, W., Joloba, M., Boom, W.H., Basu, I., Bower, J., Saraiva, M., Vasconcellos, S.E.G., Suffys, P., Koch, A., Wilkinson, R., Gail-Bekker, L., Malla, B., Ley, S.D., Beck, H.-P., de Jong, B. C., Toit, K., Sanchez-Padilla, E., Bonnet, M., Gil-Brusola, A., Frank, M., Penlap Beng, V.N., Eisenach, K., Alani, I., Ndung’u, P.W., Revathi, G., Gehre, F., Akter, S., Ntoumi, F., Stewart-Isherwood, L., Ntinginya, N.E., Rachow, A., Hoelscher, M., Cirillo, D.M., Skenders, G., Hoffner, S., Bakonyte, D., Stakenas, P., Diel, R., Crudu, V., Moldovan, O., Al-Hajoj, S., Otero, L., Barletta, F., Carter, E.J., Diero, L., Supply, P., Comas, I., Niemann, S., Gagneux, S., 2016. Mycobacterium tuberculosis lineage 4 comprises globally distributed and geographically restricted sublineages. Nat. Genet. 48, 1535–1543. https://doi.org/10.1038/ng.3704

Sudmant, P.H., Gardner, E.J., Handsaker, R.E., Abyzov, A., Huddleston, J., Zhang, Y., Ye, K., Jun, G., Hsi-Yang Fritz, M., Konkel, M.K., Malhotra, A., Stütz, A.M., Shi, X., Paolo Casale, F., Chen, J., Hormozdiari, F., Dayama, G., Chen, K., Malig, M., Chaisson, M.J.P., Walter, K., Meiers, S., Kashin, S., Garrison, E., Auton, A., Lam, H.Y.K., Jasmine Mu, X., Alkan, C., Antaki, D., Bae, T., Cerveira, E., Chines, P., Chong, Z., Clarke, L., Dal, E., Ding, L., Emery, S., Fan, X., Gujral, M., Kahveci, F., Kidd, J.M., Kong, Y., Lameijer, E.-W., McCarthy, S., Flicek, P., Gibbs, R.A., Marth, G., Mason, C. E., Menelaou, A., Muzny, D.M., Nelson, B.J., Noor, A., Parrish, N.F., Pendleton, M., Quitadamo, A., Raeder, B., Schadt, E.E., Romanovitch, M., Schlattl, A., Sebra, R., Shabalin, A.A., Untergasser, A., Walker, J.A., Wang, M., Yu, F., Zhang, C., Zhang, J., Zheng-Bradley, X., Zhou, W., Zichner, T., Sebat, J., Batzer, M.A., McCarroll, S.A., Mills, R.E., Gerstein, M.B., Bashir, A., Stegle, O., Devine, S.E., Lee, C., Eichler, E.E., Korbel, J.O., 2015. An integrated map of structural variation in 2,504 human genomes. Nature 526, 75–81. https://doi.org/10.1038/nature15394

Supply, P., Allix, C., Lesjean, S., Cardoso-Oelemann, M., Rusch-Gerdes, S., Willery, E., Savine, E., de Haas, P., van Deutekom, H., Roring, S., Bifani, P., Kurepina, N., Kreiswirth, B., Sola, C., Rastogi, N., Vatin, V., Gutierrez, M.C., Fauville, M., Niemann, S., Skuce, R., Kremer, K., Locht, C., van Soolingen, D., 2006. Proposal for Standardization of Optimized Mycobacterial Interspersed Repetitive Unit-Variable-Number Tandem Repeat Typing of Mycobacterium tuberculosis. J. Clin. Microbiol. 44, 4498–4510. https://doi.org/10.1128/JCM.01392-06

The International HapMap Consortium, 2005. A haplotype map of the human genome. Nature 437, 1299–1320. https://doi.org/10.1038/nature04226

Thye, T., Owusu-Dabo, E., Vannberg, F.O., Van Crevel, R., Curtis, J., Sahiratmadja, E., Balabanova, Y., Ehmen, C., Muntau, B., Ruge, G., Sievertsen, J., Gyapong, J., Nikolayevskyy, V., Hill, P.C., Sirugo, G., Drobniewski, F., Van De Vosse, E., Newport, M., Alisjahbana, B., Nejentsev, S., Ottenhoff, T.H.M., Hill, A.V.S., Horstmann, R.D., Meyer, C.G., 2012. Common variants at 11p13 are associated with susceptibility to tuberculosis. Nat. Genet. 44, 257–259. https://doi.org/10.1038/ng.1080

Thye, T., Vannberg, F.O., Wong, S.H., Owusu-Dabo, E., Osei, I., Gyapong, J., Sirugo, G., Sisay-Joof, F., Enimil, A., Chinbuah, M.A., Floyd, S., Warndorff, D.K., Sichali, L., Malema, S., Crampin, A.C., Ngwira, B., Teo, Y.Y., Small, K., Rockett, K., Kwiatkowski, D., Fine, P.E., Hill, P.C., Newport, M., Lienhardt, C., Adegbola, R.A., Corrah, T., Ziegler, A., Morris, A.P., Meyer, C.G., Horstmann, R.D., Hill, A.V.S., 2010. Genome-wide association analyses identifies a susceptibility locus for tuberculosis on chromosome 18q11.2. Nat. Genet. 42, 739–741. https://doi.org/10.1038/ng.639

van der Spuy, G.D., Kremer, K., Ndabambi, S.L., Beyers, N., Dunbar, R., Marais, B.J., van Helden, P.D., Warren, R.M., 2009. Changing Mycobacterium tuberculosis population highlights clade-specific pathogenic characteristics. Tuberculosis 89, 120–125. https://doi.org/10.1016/j.tube.2008.09.003

WHO, 2001. World Medical Association Declaration of Helsinki.

Yim, J.J., Selvaraj, P., 2010. Genetic susceptibility in tuberculosis. Respirology 15, 241–256. https://doi.org/10.1111/j.1440-1843.2009.01690.x

